# Machine Learning and Probabilistic Approaches for Forecasting Infectious Disease Transmission and Cases

**DOI:** 10.1101/2025.06.24.25330210

**Authors:** Md Sakhawat Hossain, Ravi Goyal, Natasha K Martin, Victor DeGruttola, Tanvir Ahammed, Christopher McMahan, Lior Rennert

**Affiliations:** Department of Public Health Sciences, Clemson University, Clemson, SC, USA; Center for Public Health Modeling and Response, Clemson University, Clemson, SC, USA; Division of Infectious Diseases & Global Public Health, University of California San Diego, La Jolla, CA, USA; Division of Biostatistics, Herbert Wertheim School of Public Health and Longevity Science, University of California San Diego, San Diego, California, USA; School of Mathematical and Statistical Sciences, Clemson University, Clemson, SC, USA

**Keywords:** Effective reproductive number, Infectious disease modeling, Machine learning, Forecasting, COVID-19

## Abstract

**Objectives:** Forecasting the effective reproductive number (*R*_*t*_) and infection case counts is critical for guiding public health responses. We developed a machine learning and probabilistic forecasting framework to predict *R*_*t*_ and daily COVID-19 cases, respectively, across South Carolina counties, with the flexibility to generalize to other infectious diseases.

**Methods:** We first estimated *R*_*t*_ using the EpiNow2 R package, which incorporates Bayesian time-series modeling and accounts for reporting delay and incubation period. These initial estimates were refined using spatial covariate-adjusted smoothing through the Integrated Nested Laplace Approximation (INLA). We then generated *R*_*t*_ forecasts using an ensemble of linear regression, random forest, and XGBoost models. Daily case forecasts were obtained by linking *R*_*t*_ trajectories with historical case data via a Poisson model.

**Results:** This ensemble-based approach outperformed EpiNow2 across different forecast horizons (7-day, 14-day, and 21-day). In the first forecast period (November 11, 2020 – February 02, 2021), the ensemble achieved a median PA of 96.5% (IQR: 95.4% – 97.1%) for 7-day horizon *R*_*t*_ forecast, compared to 87.0% (IQR: 84.4% – 89.4%) from EpiNow2. In the second period (December 11, 2022 – March 04, 2023), the ensemble attained a 93.0% median PA for *R*_*t*_ forecast (IQR: 90.8% – 95.4%), while EpiNow2 reached 86.8% (IQR: 82.5% – 89.2%). Similar trends were observed for case forecasts, with the ensemble model demonstrating improved performance.

**Conclusion:** This study presents a flexible forecasting framework that integrates Bayesian estimation, spatial smoothing, and ensemble machine learning to improve the accuracy of COVID-19 transmission and case forecasts. The approach enhances epidemic forecasting performance and offers scalable tools to support data-driven public health preparedness and response.

## 1 Introduction

The COVID-19 pandemic has significantly reshaped global priorities in public health, focusing on the importance of data-driven models to predict and manage disease spread. A crucial parameter in understanding and forecasting infectious diseases is the effective reproductive number (*R*_*t*_), which indicates the average number of secondary cases caused by an infected individual at a given time *t*. This parameter is widely utilized to monitor and assess outbreak progression and has played a critical role in informing public health interventions and policy decisions during the COVID-19 pandemic [1,2]. Values greater than 1 indicate disease transmission is increasing, while values below 1 indicate the disease is contracting [3,4].

Accurately estimating and forecasting *R*_*t*_ is crucial for assessing the transmissibility of infectious disease and guiding public health interventions, including social distancing, mask mandates, vaccination strategies, and resource allocation [1,5–8]. The *R*_*t*_ estimates can be linked to changes in policy, population behavior and immunity, pathogen evolution, and other factors [1,8–10].

Various methods have been developed to estimate R_t_, including those capable of real-time monitoring, each with its unique strengths and limitations [1,5,11–18]. EpiNow2 enhanced estimation accuracy by incorporating generation time intervals, incubation period, and reporting delays [1,11]. Most methods focus on retrospective estimation of *R*_*t*_ rather than future prediction[1,5,12,14,16,18–21]. While epidemic forecasting is a growing field, much of it focused on predicting case counts and other metrics, with relatively fewer studies explicitly forecasting the future course of *R*_*t*_ [22–26].

Machine learning has significantly advanced infectious disease modeling, aiding in outbreak detection, diagnosis, severity prediction, and forecasting of *R*_*t*_ and case counts [27–30].

Supervised learning techniques, such as random forest and gradient boosting, have been widely used for predicting disease incidence and hospitalizations, often outperforming traditional regression methods [31,32]. Combining forecasts from multiple models has emerged as a powerful approach for epidemic forecasting. Ensemble methods that combine outputs from multiple models have demonstrated improved robustness [33], particularly in scenarios with inconsistent or incomplete data [30].

In this study, we develop a framework for forecasting the effective reproductive number (R_t_) and, subsequently, daily case counts. First, we estimate *R*_*t*_ using the *R software package* EpiNow2. Next, we apply a spatial (covariate-adjusted) smoothing through INLA method to refine the *R*_*t*_ estimates [34]. We then employ regression and machine learning models to forecast *R*_*t*_ using both the initial and smooth estimates. Finally, a stochastic Poisson process is implemented to generate COVID-19 case counts based on *R*_*t*_ forecasts, utilizing historical data and the disease serial interval distribution.

## 2 Materials and Methods

We develop a forecasting framework to predict the effective reproductive number (*R*_*t*_) and daily COVID-19 case counts. We apply our method to predict these quantities for 46 South Carolina (SC) counties. *R*_*t*_ is initially estimated using the EpiNow2 *R* package, which applies Bayesian time-series modeling, accounting for reporting delay and incubation period. These estimates are then refined using a spatial (covariate-adjusted) smoothing process through the Integrated Nested Laplace Approximation (INLA) method.

To forecast *R*_*t*_, we employ linear regression (Reg), XGBoost (XGB), and Random Forest (RF) models, leveraging lagged values of the *R*_*t*_ estimates and temporal features such as the day of the year. A recursive approach is applied to generate multi-step forecasts. An ensemble-based forecasting approach is then developed by combining outputs from individual models.

For COVID-19 case forecasting, we use the forecast together with historical case data and the serial interval distribution. Forecasts are generated recursively, incorporating updated case counts at each step to refine predictions. **Figure 1** presents the workflow diagram of and daily COVID-19 case forecasts.

**Figure 1.**
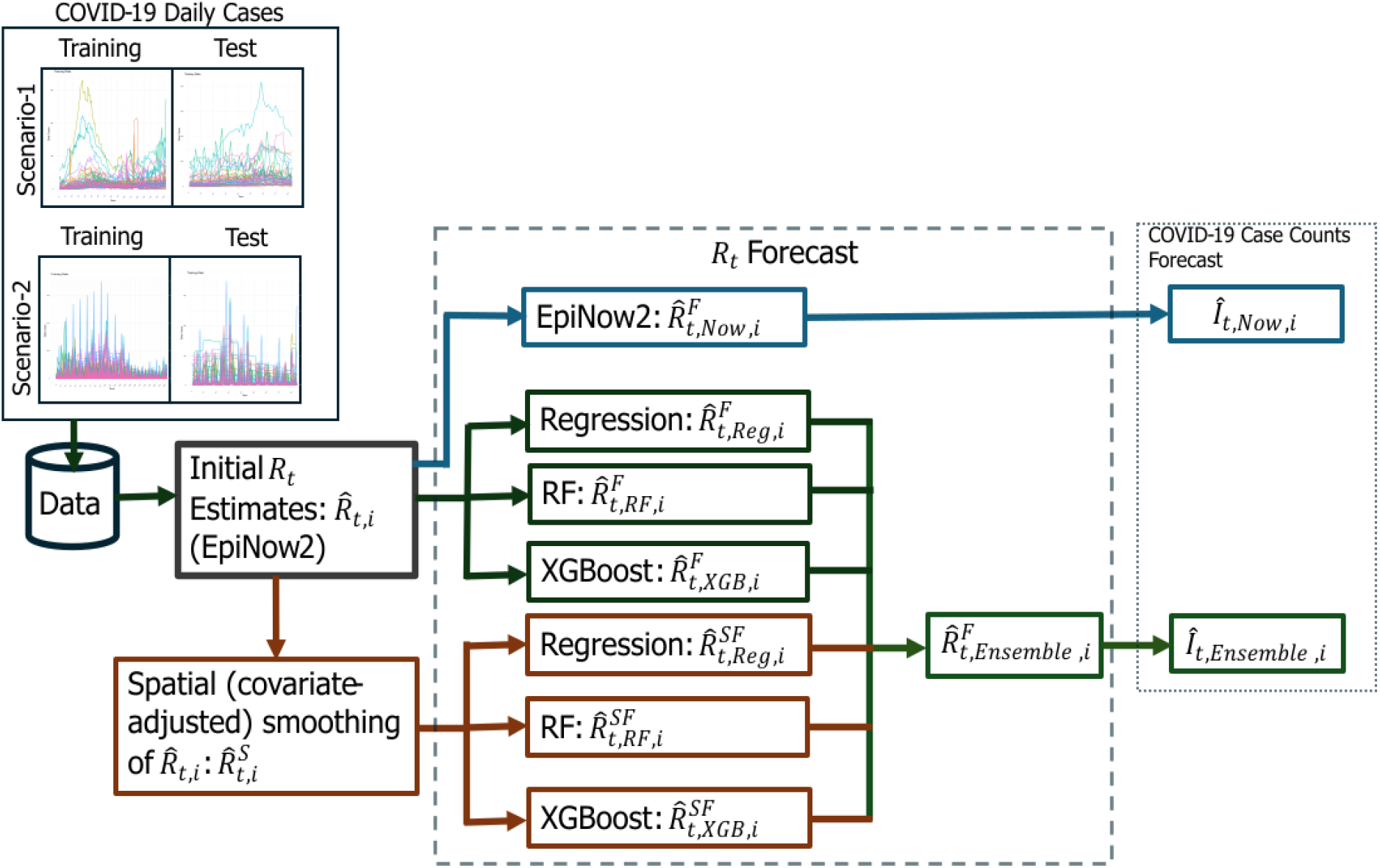
Flow diagram of the forecasting framework for effective reproductive number and COVID-19 case counts at the county level in SC. Scenario-1 includes the data from June 01, 2020, to November 10, 2020 (Training) and November 11, 2020, to February 02, 2021 (Test, forecast period), while Scenario-2 covers data from May 01, 2022, to December 10, 2022 (Training) and December 11, 2022, to March 04, 2023 (Test, forecast period). Initially, is estimated using the EpiNow2 package, followed by spatial (covariate-adjusted) smoothing via the INLA model. The initial and smoothed estimates are denoted as and, respectively. forecasting is performed using Reg, RF, and XGB on both initial and spatially smoothed estimates, followed by an ensemble-based approach for final predictions. The ensemble forecasted is denoted as 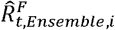, while 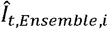 and 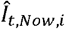 represent COVID-19 case count forecasts from the ensemble approach and EpiNow2, respectively.

We perform the initial estimation of *R*_*t*_ for the period June 01, 2020, to March 04, 2023. The forecasts for both *R*_*t*_ and COVID-19 daily case counts are generated for the time periods November 11, 2020 – February 02, 2021 (Scenario-1) and December 11, 2022 – March 04, 2023 (Scenario-2). The forecasting approach involves 7-day, 14-day, and 21-day horizon predictions over 84 days period using a rolling window method. All models are trained on *R*_*t*_ estimates from June 01, 2020, to November 10, 2020, for forecasting under Scenario-1, and from May 01, 2022, to December 10, 2022, for Scenario-2. Then we generate *R*_*t*_ forecasts through an ensemble-based approach that combines predictions from all individual models. To benchmark the performance of our models, we also produce *R*_*t*_ and case forecasts using the EpiNow2 R package, however, these forecasts are used only for comparison purposes and are not included in the ensemble.

### 2.1 Data sources

We utilize publicly available county-level COVID-19 daily case data for SC from the New York Times (NYT) GitHub repository [35]. The demographic variables, including population by age, sex, race/ethnicity, employment, and insurance coverage are used. These data are sourced from the United States Census Bureau website [36]. The social vulnerability index (SVI) is obtained from the Agency for Toxic Substances and Disease Registry website [37]. Because daily case reports show irregular updates and retrospective adjustments, we use a 7-day moving average to smooth short-term fluctuations in the data.

### 2.2 Initial estimation and spatial (covariate-adjusted) smoothing of *R*_***t***_

We first utilize the EpiNow2 R package to estimate the effective reproductive number, *R*_*t*_. It incorporates cases by reported date, generation time interval, delay distributions such as incubation and reporting delays, more details can be found at Abbott et al. [11]. The incubation period distribution and the reporting delay distribution are modeled using gamma distribution. For our analysis, we consider a mean incubation period of 5 days (95% CI, 4.94-5.06) with a standard deviation of 2.4 days [38,39], a mean serial interval distribution of 4.7 days with a standard deviation of 2.9 days [40], and a mean reporting delay of 3.2 days [41]. The initial estimate of *R*_*t*_ using EpiNow2 at a given time *t* and for a specific region *i* is denoted by 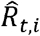. To spatially smooth *R*_*t*_ estimates, a spatial (covariate-adjusted) Bayesian model is employed, as described in [42]. The model incorporates sociodemographic covariates, including age, race, employment, insurance status, SVI, and normalized median income. We denote the smoothed estimates by 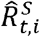.

### 2.3 Forecasting effective reproductive number (*R*_*t*_)

In this section, we outline the methodology for forecasting the effective reproductive number utilizing both estimates 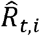 and 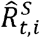 across *M* regions. These forecasts are denoted as 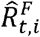 and 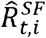 for region *i* at time *t*. For generalization, we represent these time series as *y*_*t,i*_, corresponding to either 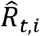 or 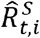. We consider the lagged estimates as predictors and incorporate temporal features, such as the day of the year (*D*_*t*_).

The time series data for region *i* is *y*_*1,i*_,*y*_*2,i*_, …, *y*_*t,i*_ where *T* is the most recent date used for forecasting. The goal is to forecast *y*_*T*+*h,i*_ for *h* ∈ {1,2 …,*H*}, where *H* is the forecasting horizon. For each region *i*, the features are constructed using *p* lagged values of the time series and the day of the year corresponding to the forecasted time point (*D*_*t*+*h*,_).

The feature matrix for all regions is constructed as below:

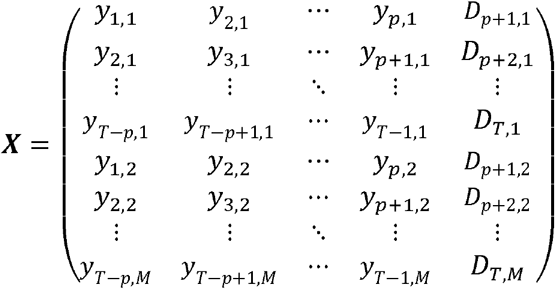

The response vector is denoted as:

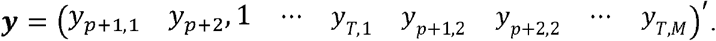

Then the model is expressed as:

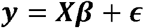

where ***y*** is response vector of (*T* − *p*) × *M* elements, **X** is features matrix of (*T* − *p*) × *M* rows and (*p* +1) columns, ***β*** is the column vector of model parameters, and *ϵ* is the residual error term.

To determine the optimal number of lagged features (*p*\*) for the final model, we train the model using values for *p* ∈ {2,3, …, *P*}. RF and XGB models are implemented in R using the ‘caret’, ‘randomForest’, and ‘xgboost’ packages. For the XGB model, we focus on tuning the learning rate (η), *L*_2_ regularization penalty (*λ*), and maximum tree depth, while for RF model, the number of trees (ntree) and the maximum number of terminal nodes (maxnodes) are tuned. All the optimizations are based on achieving the maximum PA values. After identifying the optimal lag parameter (*p*\*) and hyperparameters, the final model is trained using the data up to *y*_*T,i*_ for each region *i*. Once the final model is fitted, *h* -step horizon forecasts are generated through a recursive forecasting method.

The steps are as follows:

1. Feature construction: At time *T* + 1, form the feature vector using the last (*p*\*) observations and day of the year:

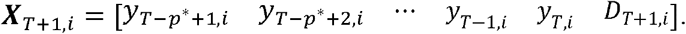
2. Prediction: Obtain the forecast 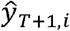 at time *T* + 1 and location *i* from the fitted Reg, RF, and XGB models.
3. Update: Append 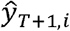 to the feature vector, discard the oldest lag, and construct

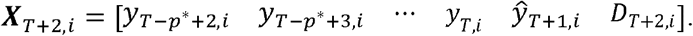
4. Iteration: Repeat steps 2-3 to generate forecasts 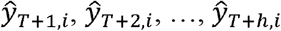 over horizon *h*.

Forecasts based on the initial estimates 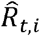 are denoted as 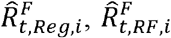, and 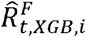, while those based on the spatially smoothed estimates 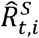 are denoted as 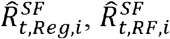, and 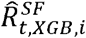. To combine these results, we implement an ensemble forecasting framework in which each model’s weight is determined from its relative percentage agreement (PA) value, with PA computed using equation (1) in Section 2.5. The ensemble forecast is denoted as 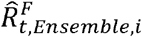.

### 2.4 Forecasting COVID-19 daily cases

To forecast the COVID-19 daily cases, we utilize a probabilistic framework that incorporates the serial interval distribution, historical case data, and forecasted effective reproductive number, 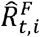. We use 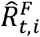 as a general term to describe the case forecasting steps; however, case forecasts are generated for each individual *R*_*t*_ forecast. The corresponding case are presented as 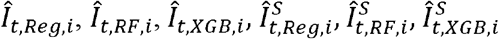 and 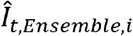.

The serial interval, representing the time between symptom onset in a primary case and secondary case, is assumed to follow a gamma distribution. This distribution is used to compute the total infectiousness (*Λ*_*t,i*_) for each day *t* at region *i*, which quantifies the potential to generate new infections from previously observed cases in s days. Total infectiousness is calculated as:

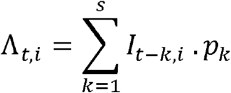

where *I*_*t*−*k,i*_ is the number of observed cases on day *t* − *k* at location *i, p*_*k*_ is the probability that a primary case generates a secondary case between *k* − 1 and days. The probabilities *p*_*k*_ represent relative probability mass function of the generation time distribution, which is typically modeled using a gamma or log-normal distribution[1,5].

Then the expected number of cases 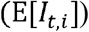 is calculated as:

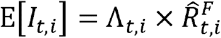

Daily case counts are then modeled as a Poisson random variable:

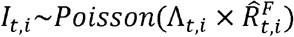

For case forecasting, we simulate full *h*-step trajectories sequentially by appending the forecasted values at each time point to the historical data before generating the next step. At each time *t*, new cases are sampled from a Poisson distribution with mean 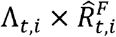, where *Λ*_*t,i*_ is computed from the updated series. This process is repeated *N* times to produce *N* forecast trajectories, and the final case forecast at each horizon is obtained by averaging across the simulated trajectories.

### 2.5 Performance evaluation of *R*_*t*_ and case forecasts

Let *o*_*t,i*_ denotes the observed values at region *i* and time *t* (e.g. daily cases, we do not know observed values for *R*_*t,i*_, use estimates 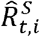), *P*_*t,i*_ represents the forecasted value at region *i* and time *t*. Then the evaluation metric is defined as follows:

Percentage Agreement (PA):

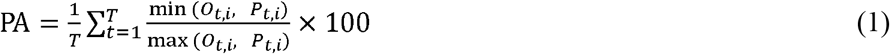

For *R*_*t*_ forecast evaluation, we compute pointwise PA, meaning that the agreement is assessed for each time point *t*, followed by averaging the PA values over the entire forecasting period. For case forecast evaluation, we first aggregate both observed and forecasted daily case counts into consecutive 7-day totals over the forecast period, calculate weekly PA by comparing aggregated forecasts with observations, and average these weekly PA values across the entire forecast period.

We use EpiNow2 R package version 1.7.1, with seed = 100 for all analyses to ensure reproducibility. We perform 5-fold cross-validation for optimal lag selection and hyperparameter tuning. A detailed summary of the selected lagged features and model-specific hyperparameters is provided in the supplementary materials (**Table S1)**.

## 3 Results

### 3.1 Evaluation of *R*_*t*_ forecasting performance

The median percentage agreement (PA) across counties for ensemble-based forecasts was higher than that of EpiNow2 (**Table 1**). For Scenario-1, the ensemble method achieved a median PA of 96.5% (IQR: 95.4% – 97.1%) across counties for 7-day horizon forecasts, while EpiNow2 reached 87.0% (IQR: 84.4% – 89.4%). Similarly, in Scenario-2, the ensemble approach maintained a median PA of 93.0% (IQR: 90.8% – 95.4%) for 7-day horizon forecasts compared to a median PA of 86.8% (IQR: 82.5% – 89.2%) for EpiNow2.

**Table 01.**
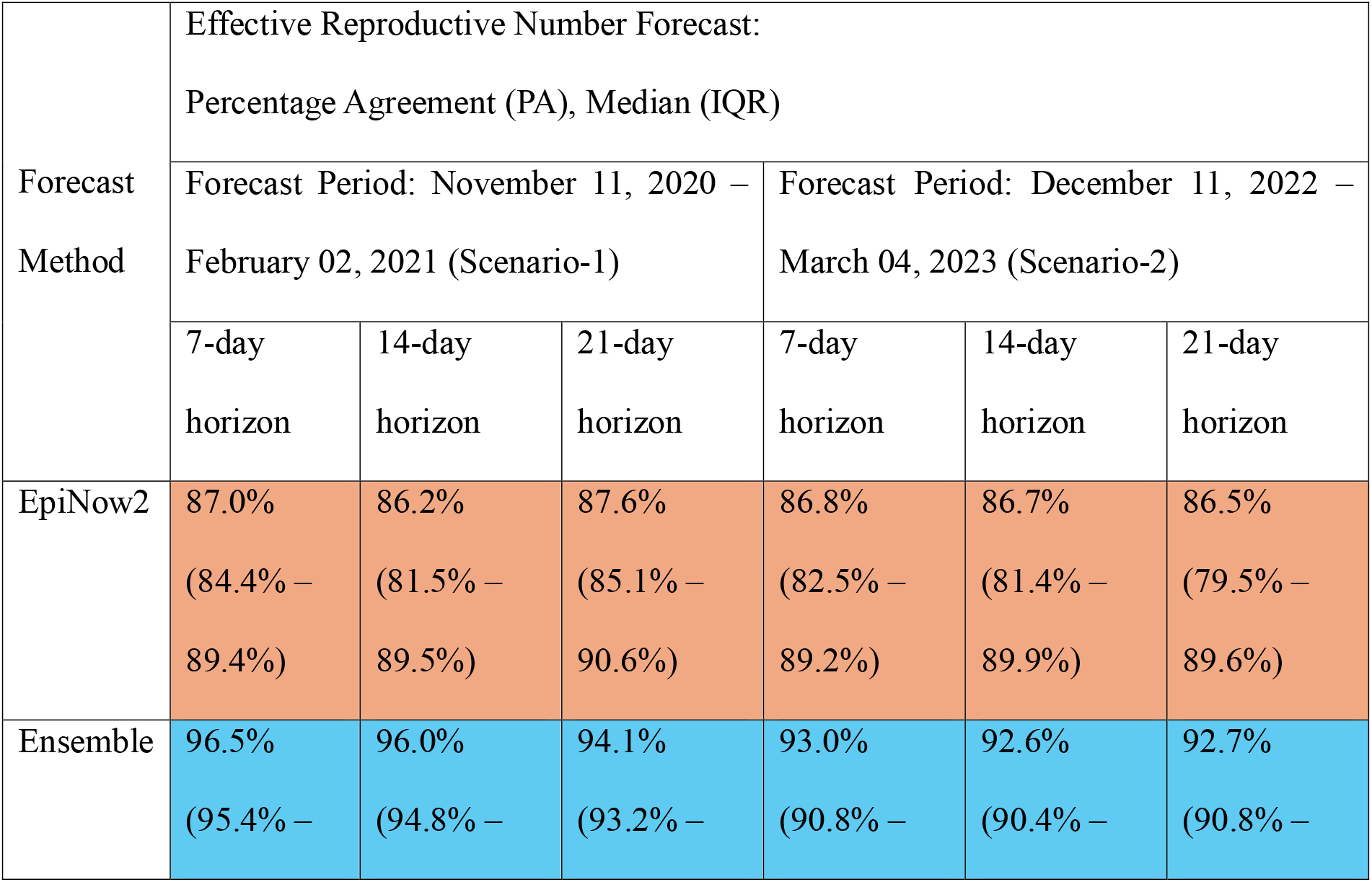
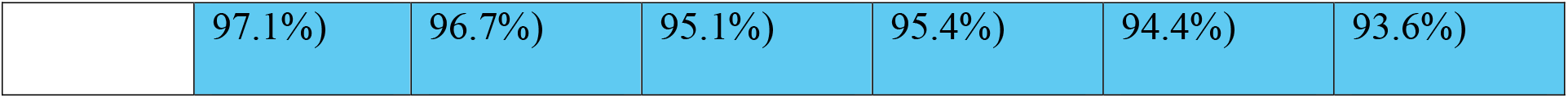
Forecast accuracy metrics for the effective reproductive number (*R*_*t*_) in SC counties during two forecast periods: November 11, 2020 – February 02, 2021 (Scenario-1) and December 11, 2022 – March 04, 2023 (Scenario-2). Forecast models were trained with data from June 01, 2020, to November 10, 2020, for Scenario-1, and from May 01, 2022, to December 10, 2022, for Scenario-2. The forecasting approach employed a rolling window method for 7-day, 14-day, and 21-day horizon predictions over 84 days period. The table presents the median PA across counties along with the IQR of county PAs.

Table 1 provides a detailed comparison of forecasting accuracy for 7-day, 14-day, and 21-day horizons of forecasts for 84 days period during both Scenario-1 and Scenario-2. The ensemble-based approach outperformed EpiNow2 across all forecast horizons. The relative difference in the 7-day horizon forecast PA was 10.9% higher for the ensemble approach in Scenario-1 (96.5% vs 87.0%) and 7.1% higher in Scenario-2 (93.0% vs. 86.8%). The relative difference in 21-day horizon PA was 7.4% higher for the ensemble approach in Scenario-1 (94.1% vs 87.6%) and 7.2% higher in Scenario-2 (92.7% vs 86.5%).

In counties with higher COVID-19 case counts (**Figure 2**, left panel), both methods captured trends well, but EpiNow2 (red lines) exhibited more fluctuations, particularly in Horry and Lexington. In counties with lower COVID-19 case counts (**Figure 2**, right panel), the ensemble approach still performed better.

**Figure 2.**
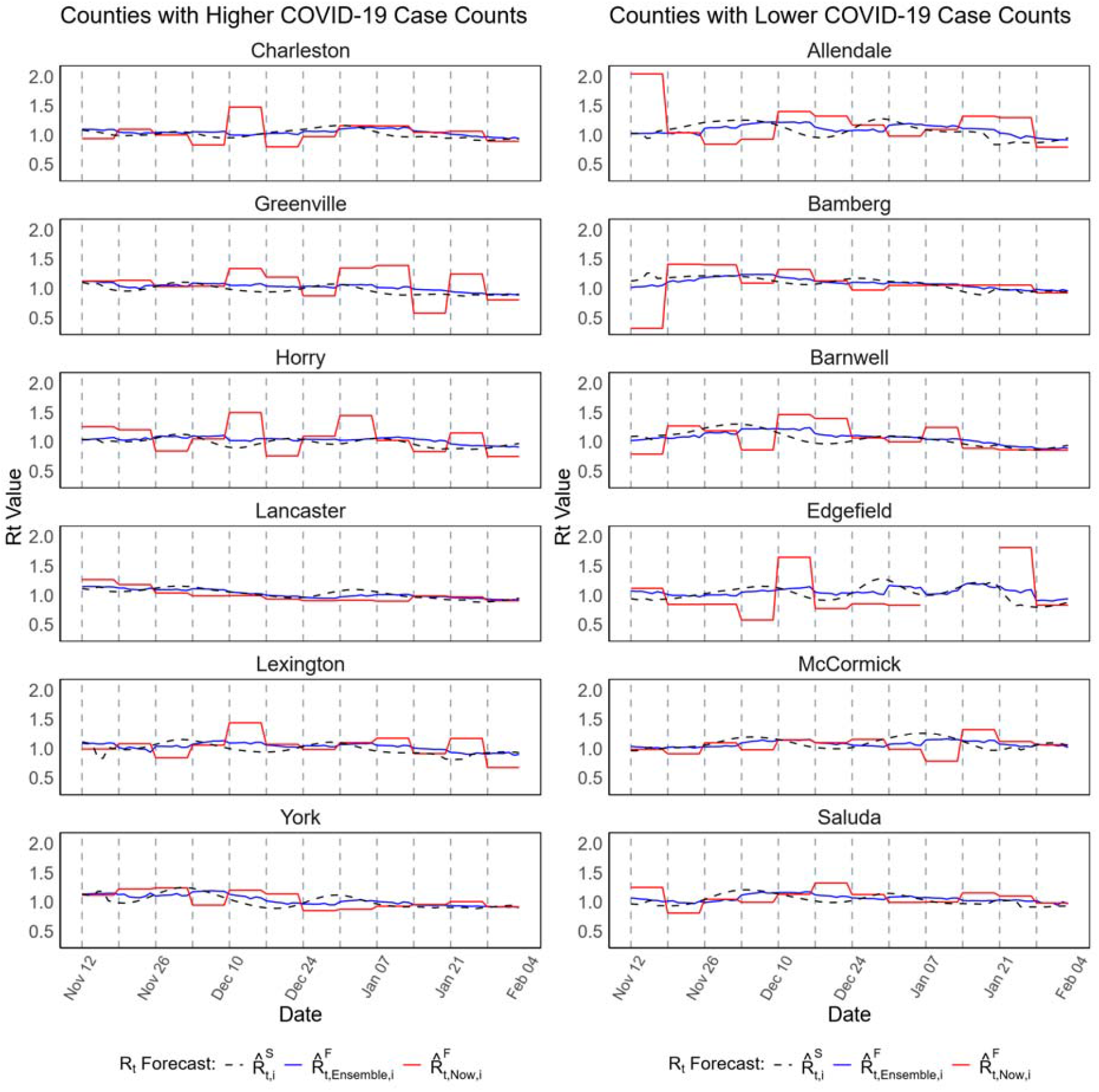
Forecast of at the county level in SC during Scenario-1 (November 11, 2020 – February 02, 2021). The left panel presents forecasts for six counties with higher COVID-19 case counts, while the right panel displays forecasts for six counties with lower case counts. The plots compare the ensemble-based forecasts (blue lines) with those generated using the **EpiNow2** R package (red lines), alongside the spatially (covariate-adjusted) smoothed estimates, (black dashed lines), where presents the county, denotes the time point (day), and “Now refers to the **EpiNow2** method. Forecasts were generated using a rolling window approach for 7-day horizon predictions over 84 days period.

Across all counties, the ensemble approach provided higher agreement with observed trends, while EpiNow2 showed more variability, particularly in some counties with significant fluctuations (e.g., Horry and Edgefield). The supplementary materials provide further insights into forecast performance across all counties. **Figure S1** presents 7-day horizon *R*_*t*_ forecasts for all 46 counties, while **Figures S2** and **S3** display the 14-day and 21-day horizon forecasts, respectively, for Scenario-1. Similarly, **Figures S4**-**S6** provide the 7-day, 14-day, and 21-day forecasts for Scenario-2, reaffirming that the ensemble model outperformed EpiNow2.

Figure S7 presents the 7-day horizon forecasts of *R*_*t*_ for all 46 counties in SC under Scenario-1, where we compared the spatially smoothed forecast from EpiNow2 with the ensemble-based forecasts. Alongside the spatially smooth *R*_*t*_ estimates. **Figure S8** compares the forcasted *R*_*t*_ values with the initial *R*_*t*_ estimates, allowing assessment of model performance without spatial smoothing. **Table S2** summarizes the corresponding forecast accuracies for both Scenario-1 and Scenario-2. Across all horizons and both scenarios, the ensemble forecasts outperformed EpiNow2. For example, the ensemble achieved a median PA of 96.2% for 7-day horizon forecasts in Scenario-1 and 92.2% in Scenario-2. These correspond to approximately 12.3% and 4.1% higher relative performance compared to EpiNow2 forecasts, respectively.

### 3.2 Evaluation of COVID-19 case forecasting performance

The results indicated that the ensemble approach outperformed EpiNow2, particularly for short-term forecasts (7-day and 14-day horizons). In Scenario-1, the ensemble model achieved a median PA of 87.3% (IQR: 84.0% – 88.8%) for 7-day forecasts, slightly outperforming EpiNow2, which had a median PA of 83.8% (IQR: 81.6% – 86.9%). For longer horizons (21-day forecasts), the ensemble approach maintained a median PA of 82.6% (IQR: 80.0% – 86.5%), while EpiNow2 showed a lower median PA of 67.6% (IQR: 61.3% – 75.1%), demonstrating that the ensemble method provided greater stability in extended forecasting periods.

A similar trend was observed in Scenario-2, where ensemble-based forecasts continued to outperform EpiNow2 across all forecast horizons. The relative improvement in PA was more evident for longer-term predictions, with the ensemble model achieving a median PA of 74.6% (IQR: 66.7% – 78.7%) for 21-day forecasts, compared to 66.2% (IQR: 57.4% – 73.2%) for EpiNow2. **Table 2** presents a detailed comparison of forecast accuracy metrics for 7-day, 14-day, and 21-day horizon predictions across both scenarios.

**Table 2.**
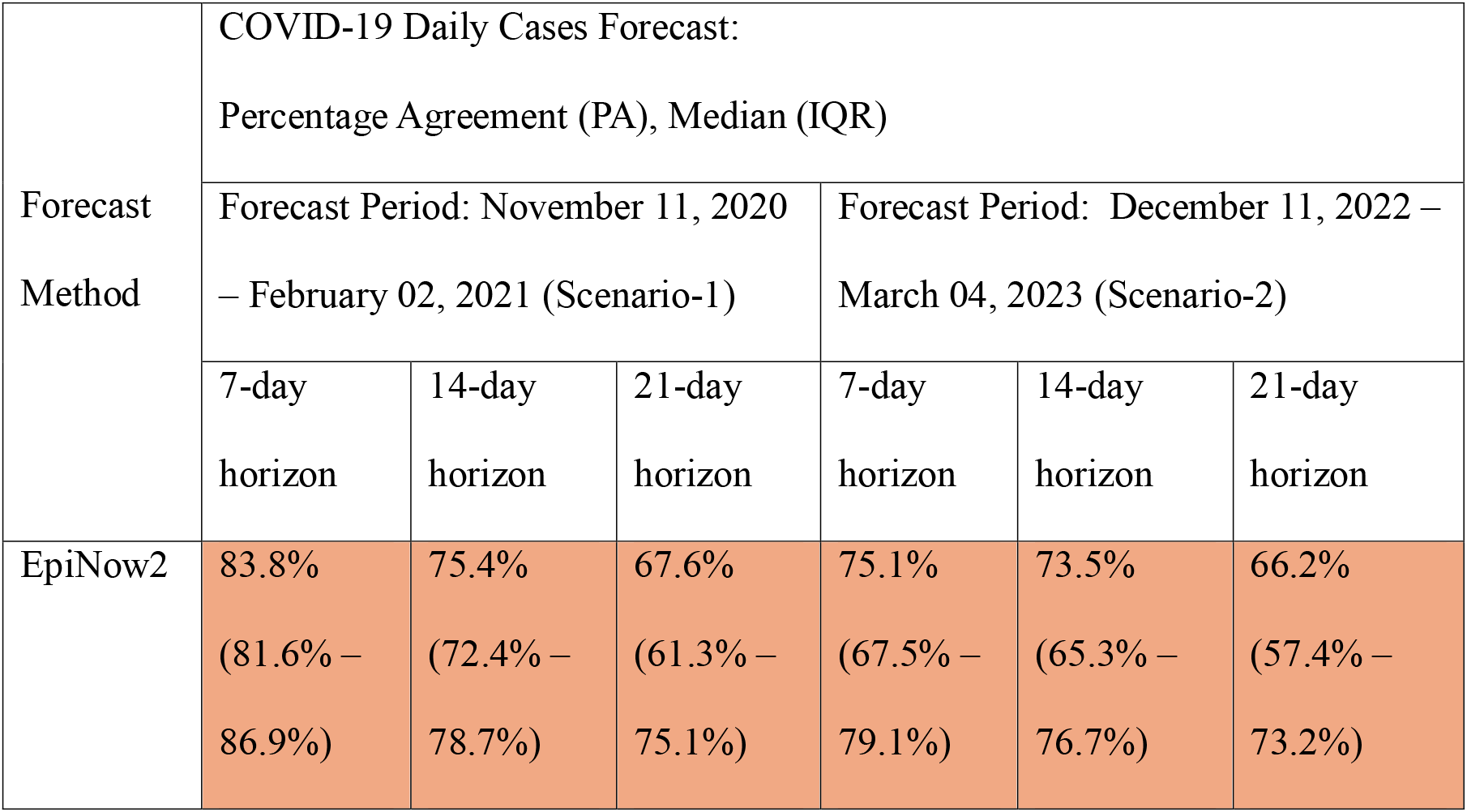
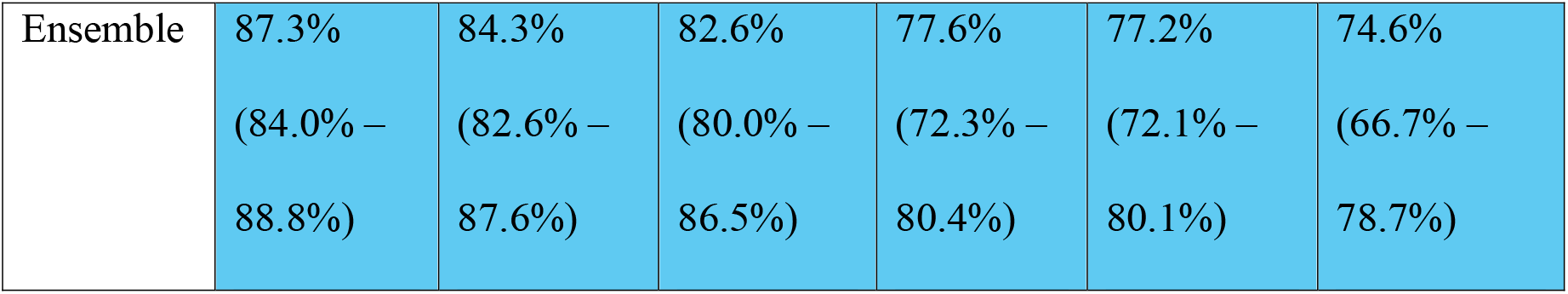
Forecast accuracy metrics for COVID-19 daily case counts at the county level in SC during Scenario-1 (November 11, 2020 – February 2, 2021) and Scenario-2 (December 11, 2022 – March 4, 2023). Forecast accuracy was evaluated using PA with median and IQR across all 46 counties. Case forecasts were generated using the EpiNow2 R package and a probabilistic Poisson model based on the ensemble-based *R*_*t*_ forecasts 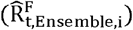. Forecasts were conducted for 7-day, 14-day, and 21-day horizon predictions over 84 days period using a rolling window approach.

The results indicate that the ensemble approach produced more stable and accurate case forecasts than EpiNow2, with differences varying by county and case count levels (**Figure 3**). In higher case count counties (Charleston, Greenville, Horry, Lancaster, Lexington, York), the ensemble model captured trends more smoothly, whereas EpiNow2 exhibited abrupt spikes and larger deviations from observed case counts, particularly in Horry and Lexington. In lower case count counties (Allendale, Bamberg, Barnwell, Edgefield, McCormick, Saluda), the performance gap between the two methods was more evident in certain areas, such as Edgefield.

**Figure 3.**
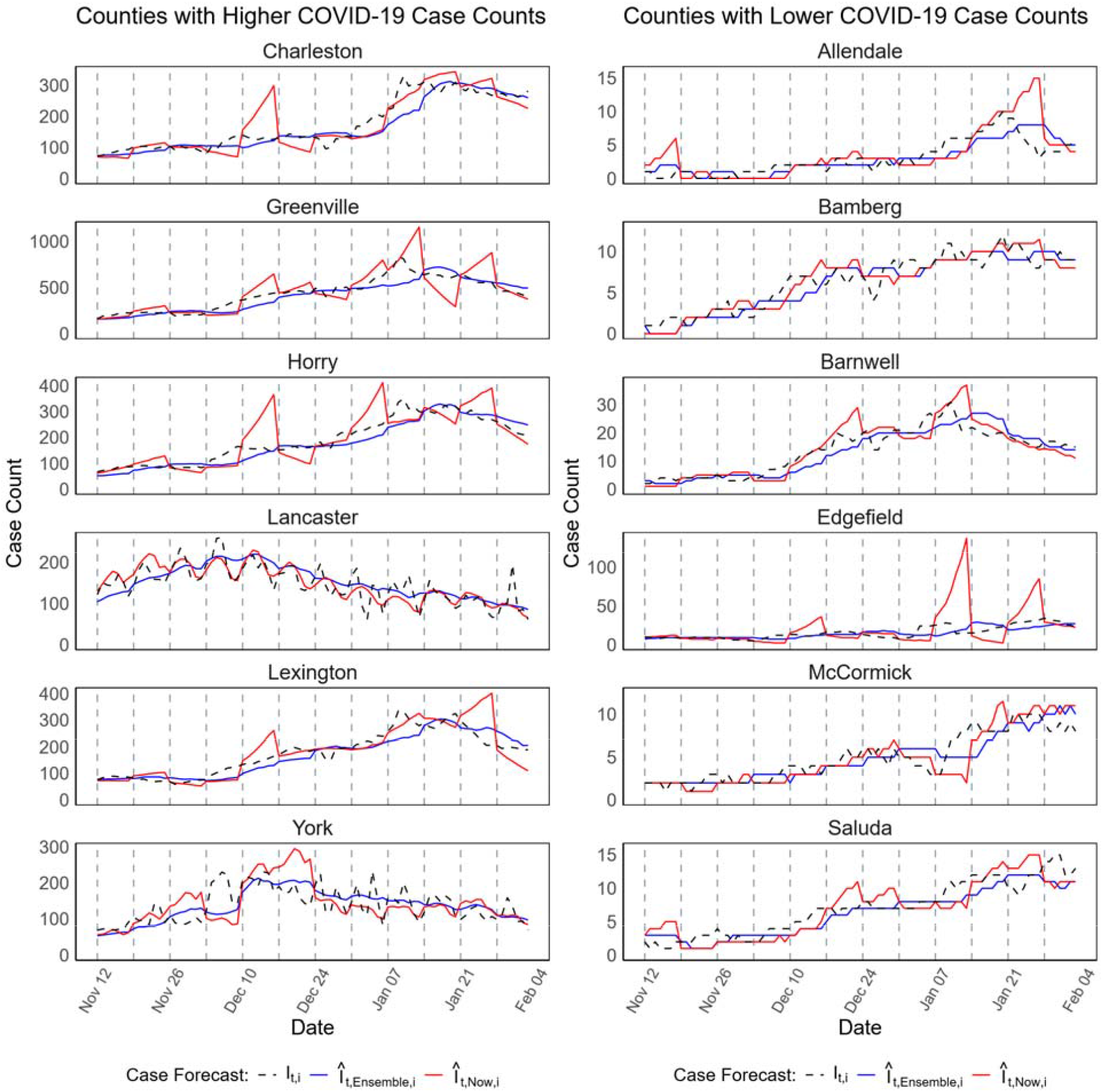
Forecast of COVID-19 case counts at the county level in SC during Scenario-1 (November 11, 2020 February 02, 2021). The left panel presents forecasts for six counties with higher COVID-19 case counts, while the right panel displays forecasts for six counties with lower case counts. The plots compare EpiNow2 forecasts (red lines) and ensemble-based forecasts (blue lines) against the observed daily case counts (black dashed lines). The forecasts were generated for 7-day horizon predictions over 84 days period using a rolling window approach.

Overall, the results confirm that the ensemble approach provided more accurate and stable case forecasts across all time horizons and geographical regions. The supplementary materials provide a comprehensive summary of forecast performance across all counties. **Figures S9**-**S11** (Scenario-1) and **Figures S12**-**S14** (Scenario-2) present case forecasts for all counties at 7-day, 14-day, and 21-day horizon, respectively.

**3.2 *R***_***t***_ **and COVID-19 case forecasting performance of individual models**

**Table S3** indicates that RF models achieved higher median PA values compared to Reg and XGB models, especially when trained on spatially smoothed estimates. For instance, during Scenario-1, XGB model with spatial smoothing attained a median PA of 97.3% (IQR: 96.8%–97.7%) for 7-day horizon forecasts, whereas Reg and RF models with spatial smoothing had median PAs of 97.3% (IQR: 96.7%–97.7%) and 96.4% (IQR: 95.6%–96.7%), respectively. EpiNow2 with spatial smoothing obtained a median PA of 87.6% (IQR: 86.6%–88.9%).

As shown in **Table S4**, the XGB model with spatial smoothing outperformed all the other individual models in forecasting daily COVID-19 cases. In Scenario-1, RF with spatial smoothing achieved a median PA of 87.7% (IQR: 84.9%–89.7%) for 7-day horizon forecast, surpassing Reg and XGB models with spatial smoothing, which had median PAs of 86.7% (IQR: 83.1%–87.8%) and 86.8% (IQR: 83.4%–87.8%), respectively.

The forecasting performance of Reg, RF, and XGB models is evaluated through **Figures S15-S20** for *R*_*t*_ forecasts and **Figures S21-S26** for COVID-19 case forecasts, across different forecast horizons. For *R*_*t*_ forecasting, all Reg, RF, and XGB models consistently provided predictions, closely following the smoothed *R*_*t*_ estimates across different counties. For COVID-19 case forecasting in Scenario-1, XGB and RF with spatial smoothing outperformed the other models, particularly in counties with complex transmission dynamics. In Scenario-2 case forecasting, XGB and RF without spatial smoothing achieved higher prediction accuracy compared to the other models.

## 4 Discussion

Accurate forecasting of *R*_*t*_ and COVID-19 case counts is critical for epidemic control and public health decision-making. Previous studies have demonstrated the importance of real-time *R*_*t*_ estimation in outbreak control, emphasizing its role in quantifying disease transmissibility and guiding policy responses [1,18]. Traditional statistical models, such as those used in Bayesian inference-based approaches like EpiNow2, have been widely employed to estimate *R*_*t*_ in real time [1,43]. This study integrates machine learning techniques and a probabilistic framework to address the challenges of forecasting disease transmission and cases. Our findings indicate that the ensemble-based approach outperformed the widely used EpiNow2 model across different forecasting horizons.

The practical implications of our forecasting framework are significant. Accurate *R*_*t*_ predictions enable early detection of outbreaks, allowing for timely interventions such as targeted lockdowns or vaccination campaigns. Similarly, reliable case forecasts inform resource allocation, ensuring that medical supplies, hospital beds, and testing infrastructure are optimally distributed. The ability to evaluate policy effectiveness through changes in *R*_*t*_ and forecasted trends supports evidence-based decision-making. Although our analysis focused on COVID-19 transmission across counties in SC, the proposed ensemble framework is generalizable to other infectious diseases and spatial scales. With appropriate input data, it can be applied to finer geographic resolutions (e.g., ZIP code or census tract levels) or to different pathogens such as influenza, or RSV to support region-specific public health decision-making.

While the proposed forecasting framework demonstrated improved accuracy compared to EpiNow2, several limitations must be acknowledged. Previous studies have shown that geographically refined predictions can better inform targeted responses, including mobile health clinic (MHC) deployments [44]. More detailed spatial forecasts would allow public health officials to allocate resources more effectively, particularly in underserved communities where healthcare access is limited. Additionally, real-time forecasting remains challenging due to potential delays in obtaining case data from state health departments. In practice, alternative data sources, such as electronic health records (EHR) from health systems, may be necessary to supplement official reports. Previous research has demonstrated promise in leveraging alternative data sources for predictive modeling, but adapting models for real-time implementation in such settings requires further validation [44]. Furthermore, forecast accuracy declines as the prediction horizon increases, a common issue in time series forecasting.

Future research could focus on integrating Bayesian deep learning techniques with real-time data streams, such as mobility, genomic surveillance, wastewater data, and electronic health records, to improve forecast accuracy and uncertainty quantification.

## Conclusion

In this study, we developed a forecasting framework to predict the effective reproductive number (*R*_*t*_) and COVID-19 daily case counts using a combination of machine learning models and probabilistic approaches. By leveraging Bayesian estimates, spatial smoothing, and recursive forecasting, our approach enhances the accuracy of epidemic predictions. The ensemble-based methodology further refines forecast by optimally combining individual model outputs using PA as a weighting criterion. Our results demonstrate that incorporating spatially adjusted estimates improves forecast performance across different pandemic waves and forecasting horizons. Future work should explore adaptive modeling techniques that integrate real-time mobility data and genomic surveillance to refine predictions further. Additionally, expanding the framework to accommodate multiple infectious diseases could enhance its applicability for broader epidemiological forecasting. Overall, this study contributes to advancing epidemic forecasting by integrating machine learning and probabilistic models in a flexible and scalable framework. These findings can support data-driven policy interventions, improve epidemic preparedness, and facilitate effective public health responses in future outbreaks.

## Data Availability

All code and public data used for analyses and figure generations are available at https://github.com/mdsakhh/Machine-Learning-and-Probabilistic-Approach-for-Rt-and-COVID-19-Case-Forecasting and from the corresponding author upon request.

## Ethics statement

Ethical review for this study was obtained by the Institutional Review Board of Clemson University (2020-0150). No consent was needed for this study.

## CRediT authorship contribution statement

LR, CM, SH conceptualized and developed the methodology for this study. SH conducted formal analysis, visualization, validation, data curation, and drafted the original manuscript with contributions from LR. LR, CM, and RG provided critical feedback and guidance throughout the research process. All authors contributed to interpretation of results. LR and CM oversaw project administration and supervision. LR acquired the funding for this study. All authors participated in reviewing and editing the manuscript.

## Acknowledgements

This project has been funded by the National Library of Medicine of the National Institutes of Health (NIH) under award number R01LM014193 and the Center for Forecasting and Outbreak Analytics of the Centers for Disease Control and Prevention (CDC) under award number NU38FT000011. The content and decision to publish is solely based on the authors of this study and does not necessarily represent the official views of the NIH or CDC. The funders had no role in the study design, data collection and analysis, decision to publish, or preparation of this manuscript.

## Declaration of generative AI and AI-assisted technologies in the writing process

During the preparation of this work the authors used GPT-4o and GPT-5 to assist with writing and grammar checks. After using these tools, the authors reviewed and edited the content as needed.

## References

1. Gostic KM, McGough L, Baskerville EB, Abbott S, Joshi K, Tedijanto C, et al. Practical considerations for measuring the effective reproductive number, Rt. PLOS Comput Biol. 2020 Dec 10;16(12):e1008409.

2. Huisman JS, Scire J, Angst DC, Li J, Neher RA, Maathuis MH, et al. Estimation and worldwide monitoring of the effective reproductive number of SARS-CoV-2. Davenport MP, editor. eLife. 2022 Aug 8;11:e71345.

3. Anderson RM, May RM. Infectious Diseases of Humans: Dynamics and Control. Oxford, New York: Oxford University Press; 1992. 766 p.

4. Nishiura H, Chowell G. The Effective Reproduction Number as a Prelude to Statistical Estimation of Time-Dependent Epidemic Trends. In: Chowell G, Hyman JM, Bettencourt LMA, Castillo-Chavez C, editors. Mathematical and Statistical Estimation Approaches in Epidemiology [Internet]. Dordrecht: Springer Netherlands; 2009 [cited 2023 Dec 19]. p. 103–21. Available from: http://link.springer.com/10.1007/978-90-481-2313-1_5

5. Cori A, Ferguson NM, Fraser C, Cauchemez S. A new framework and software to estimate time-varying reproduction numbers during epidemics. Am J Epidemiol. 2013;178(9):1505– 12.

6. Flaxman S, Mishra S, Gandy A, Unwin HJT, Mellan TA, Coupland H, et al. Estimating the effects of non-pharmaceutical interventions on COVID-19 in Europe. Nature. 2020 Aug;584(7820):257–61.

7. Fraser C. Estimating Individual and Household Reproduction Numbers in an Emerging Epidemic. PLOS ONE. 2007 Aug 22;2(8):e758.

8. Kucharski AJ, Russell TW, Diamond C, Liu Y, Edmunds J, Funk S, et al. Early dynamics of transmission and control of COVID-19: a mathematical modelling study. Lancet Infect Dis. 2020 May;20(5):553–8.

9. Banholzer N, Feuerriegel S, Vach W. Estimating and explaining cross-country variation in the effectiveness of non-pharmaceutical interventions during COVID-19. Sci Rep. 2022 May 9;12(1):7526.

10. Lison A, Persson J, Banholzer N, Feuerriegel S. Estimating the effect of mobility on SARS-CoV-2 transmission during the first and second wave of the COVID-19 epidemic, Switzerland, March to December 2020. Eurosurveillance. 2022 Mar 10;27(10):2100374.

11. Abbott S, Hellewell J, Sherratt K, Gostic K, Hickson J, Badr HS, et al. EpiNow2: estimate real-time case counts and time-varying epidemiological parameters. R Package Version 01 0 [Internet]. 2020 [cited 2024 Feb 13]; Available from: https://scholar.google.com/scholar?cluster=1308097242765034406&hl=en&oi=scholarr

12. Wallinga J, Teunis P. Different Epidemic Curves for Severe Acute Respiratory Syndrome Reveal Similar Impacts of Control Measures. Am J Epidemiol. 2004 Sep 15;160(6):509– 16.

13. Meyer S, Held L, Höhle M. Spatio-Temporal Analysis of Epidemic Phenomena Using the R Package surveillance. J Stat Softw [Internet]. 2017 [cited 2024 Mar 6];77(11). Available from: http://arxiv.org/abs/1411.0416

14. Bettencourt LM, Ribeiro RM. Real time bayesian estimation of the epidemic potential of emerging infectious diseases. PloS One. 2008;3(5):e2185.

15. Cauchemez S, Boëlle PY, Donnelly CA, Ferguson NM, Thomas G, Leung GM, et al. Real-time estimates in early detection of SARS. Emerg Infect Dis. 2006;12(1):110.

16. Thompson RN, Stockwin JE, van Gaalen RD, Polonsky JA, Kamvar ZN, Demarsh PA, et al. Improved inference of time-varying reproduction numbers during infectious disease outbreaks. Epidemics. 2019 Dec 1;29:100356.

17. Wallinga J, Lipsitch M. How generation intervals shape the relationship between growth rates and reproductive numbers. Proc R Soc B Biol Sci. 2006 Nov 28;274(1609):599–604.

18. Parag KV. Improved estimation of time-varying reproduction numbers at low case incidence and between epidemic waves. PLOS Comput Biol. 2021 Sep 7;17(9):e1009347.

19. Bokányi E, Vizi Z, Koltai J, Röst G, Karsai M. Real-time estimation of the effective reproduction number of COVID-19 from behavioral data. Sci Rep. 2023 Dec 5;13(1):21452.

20. Nash RK, Nouvellet P, Cori A. Real-time estimation of the epidemic reproduction number: Scoping review of the applications and challenges. PLOS Digit Health. 2022 Jun 27;1(6):e0000052.

21. Won YS, Son WS, Choi S, Kim JH. Estimating the instantaneous reproduction number (Rt) by using particle filter. Infect Dis Model. 2023 Dec 1;8(4):1002–14.

22. Mellor J, Tang ML, Jones O, Ward T, Riley S, Deeny SR. Forecasting COVID-19, influenza, and RSV hospitalizations over winter 2023–4 in England. Int J Epidemiol. 2025 Jun 1;54(3):dyaf066.

23. Dixon S, Keshavamurthy R, Farber DH, Stevens A, Pazdernik KT, Charles LE. A Comparison of Infectious Disease Forecasting Methods across Locations, Diseases, and Time. Pathogens. 2022 Jan 29;11(2):185.

24. Tang CY, Gao C, Prasai K, Li T, Dash S, McElroy JA, et al. Prediction models for COVID-19 disease outcomes. Emerg Microbes Infect. 13(1):2361791.

25. Gibson GC, Moran KR, Reich NG, Osthus D. Improving probabilistic infectious disease forecasting through coherence. PLOS Comput Biol. 2021 Jan 6;17(1):e1007623.

26. Lutz CS, Huynh MP, Schroeder M, Anyatonwu S, Dahlgren FS, Danyluk G, et al. Applying infectious disease forecasting to public health: a path forward using influenza forecasting examples. BMC Public Health. 2019 Dec 10;19(1):1659.

27. Gao Y, Cai GY, Fang W, Li HY, Wang SY, Chen L, et al. Machine learning based early warning system enables accurate mortality risk prediction for COVID-19. Nat Commun. 2020 Oct 6;11(1):5033.

28. Mei X, Lee HC, Diao K yue, Huang M, Lin B, Liu C, et al. Artificial intelligence–enabled rapid diagnosis of patients with COVID-19. Nat Med. 2020 Aug;26(8):1224–8.

29. Wynants L, Calster BV, Collins GS, Riley RD, Heinze G, Schuit E, et al. Prediction models for diagnosis and prognosis of covid-19: systematic review and critical appraisal. BMJ. 2020 Apr 7;369:m1328.

30. Arik S, Li CL, Yoon J, Sinha R, Epshteyn A, Le L, et al. Interpretable Sequence Learning for Covid-19 Forecasting. In: Advances in Neural Information Processing Systems [Internet]. Curran Associates, Inc.; 2020 [cited 2023 Nov 7]. p. 18807–18. Available from: https://proceedings.neurips.cc/paper/2020/hash/d9dbc51dc534921589adf460c85cd824-Abstract.html

31. Fife DA, D’Onofrio J. Common, uncommon, and novel applications of random forest in psychological research. Behav Res Methods. 2023 Aug 1;55(5):2447–66.

32. King C, Strumpf E. Applying random forest in a health administrative data context: a conceptual guide. Health Serv Outcomes Res Methodol. 2022 Mar 1;22(1):96–117.

33. Oidtman RJ, Omodei E, Kraemer MUG, Castañeda-Orjuela CA, Cruz-Rivera E, Misnaza-Castrillón S, et al. Trade-offs between individual and ensemble forecasts of an emerging infectious disease. Nat Commun. 2021;12(1).

34. Hossain MS, Goyal R, Martin NK, DeGruttola V, Chowdhury MM, McMahan C, et al. A flexible framework for local-level estimation of the effective reproductive number in geographic regions with sparse data. BMC Med Res Methodol. 2025 Mar 18;25(1):73.

35. The New York Times. Coronavirus (Covid-19) Data in the United States [Internet]. 2021 [cited 2023 Oct 6]. Available from: https://github.com/nytimes/covid-19-data

36. U.S. Census Bureau. Census Data [Internet]. 2020. Available from: https://data.census.gov/profile

37. Agency for Toxic Substances and Disease Registry. CDC/ATSDR SVI: Data and Documentation Download [Internet]. 2022. Available from: https://www.atsdr.cdc.gov/placeandhealth/svi/data_documentation_download.html

38. Grant R, Charmet T, Schaeffer L, Galmiche S, Madec Y, Platen CV, et al. Impact of SARS-CoV-2 Delta variant on incubation, transmission settings and vaccine effectiveness: Results from a nationwide case-control study in France. Lancet Reg Health – Eur [Internet]. 2022 Feb 1 [cited 2024 May 10];13. Available from: https://www.thelancet.com/journals/lanepe/article/PIIS2666-7762(21)00264-7/fulltext

39. Lauer SA, Grantz KH, Bi Q, Jones FK, Zheng Q, Meredith HR, et al. The Incubation Period of Coronavirus Disease 2019 (COVID-19) From Publicly Reported Confirmed Cases: Estimation and Application. Ann Intern Med. 2020 May 5;172(9):577–82.

40. Nishiura H, Linton NM, Akhmetzhanov AR. Serial interval of novel coronavirus (COVID-19) infections. Int J Infect Dis IJID Off Publ Int Soc Infect Dis. 2020 Apr;93:284–6.

41. Harris JE. Timely epidemic monitoring in the presence of reporting delays: anticipating the COVID-19 surge in New York City, September 2020. BMC Public Health. 2022 May 2;22(1):871.

42. Hossain MS, Goyal R, Martin NK, DeGruttola V, Chowdhury MM, McMahan C, et al. A Flexible Framework for Local-Level Estimation of the Effective Reproductive Number in Geographic Regions with Sparse Data [Internet]. medRxiv; 2024 [cited 2024 Dec 12]. p. 2024.11.06.24316859. Available from: https://www.medrxiv.org/content/10.1101/2024.11.06.24316859v1

43. Abbott S, Lison A, Funk S. epinowcast: Flexible hierarchical nowcasting. Zenodo [Internet]. 2021 [cited 2024 Feb 13]; Available from: https://samabbott.co.uk/presentations/2023/royal-society-epinowcast.pdf

44. Gezer F, Howard KA, Litwin AH, Martin NK, Rennert L. Identification of factors associated with opioid-related and hepatitis C virus-related hospitalisations at the ZIP code area level in the USA: an ecological and modelling study. Lancet Public Health. 2024 Jun 1;9(6):e354–64.

