## Supplementary Information 1 for "Machine Learning and Probabilistic Approaches for Forecasting Infectious Disease Transmission and Cases"

**Supplementary Materials:**

**Table S1*.*** Summary of optimal lag (lagged feature) selection and hyperparameter tuning for Reg, RF, and XGB models in Scenario-1 (November 11, 2020 – February 02, 2021) and Scenario-2 (December 11, 2022 – March 04, 2023), based on 5-fold cross-validation.

| **Forecast**  **Period** | **Parameters** | **Regression** | **Regression (Smooth)** | **RF** | **RF (Smooth)** | **XGBoost** | **XGBoost (Smooth)** |
| --- | --- | --- | --- | --- | --- | --- | --- |
| **Scenario-1** | optimal lag | 12 | 4 | 10 | 3 | 18 | 7 |
|  | $L_{2}$ penalty $(\lambda)$ | --- | --- | --- | --- | 3 | 5 |
|  | learning rate $(\eta)$ | --- | --- | --- | --- | 0.05 | 0.05 |
|  | max depth | --- | --- | --- | --- | 8 | 10 |
|  | node size | --- | --- | 10 | 10 | --- | --- |
|  | ntree | --- | --- | 500 | 400 | --- | --- |
|  | maxnodes | --- | --- | 150 | 150 | --- | --- |
| **Scenario-2** | optimal lag | 19 | 19 | 3 | 3 | 14 | 24 |
|  | $L_{2}$ penalty $(\lambda)$ | --- | --- | --- | --- | 2 | 1 |
|  | learning rate $(\eta)$ | --- | --- | --- | --- | 0.05 | 0.05 |
|  | max depth | --- | --- | --- | --- | 8 | 8 |
|  | node size | --- | --- | 10 | 10 | --- | --- |
|  | ntree | --- | --- | 200 | 500 | --- | --- |
|  | maxnodes | --- | --- | 150 | 150 | --- | --- |

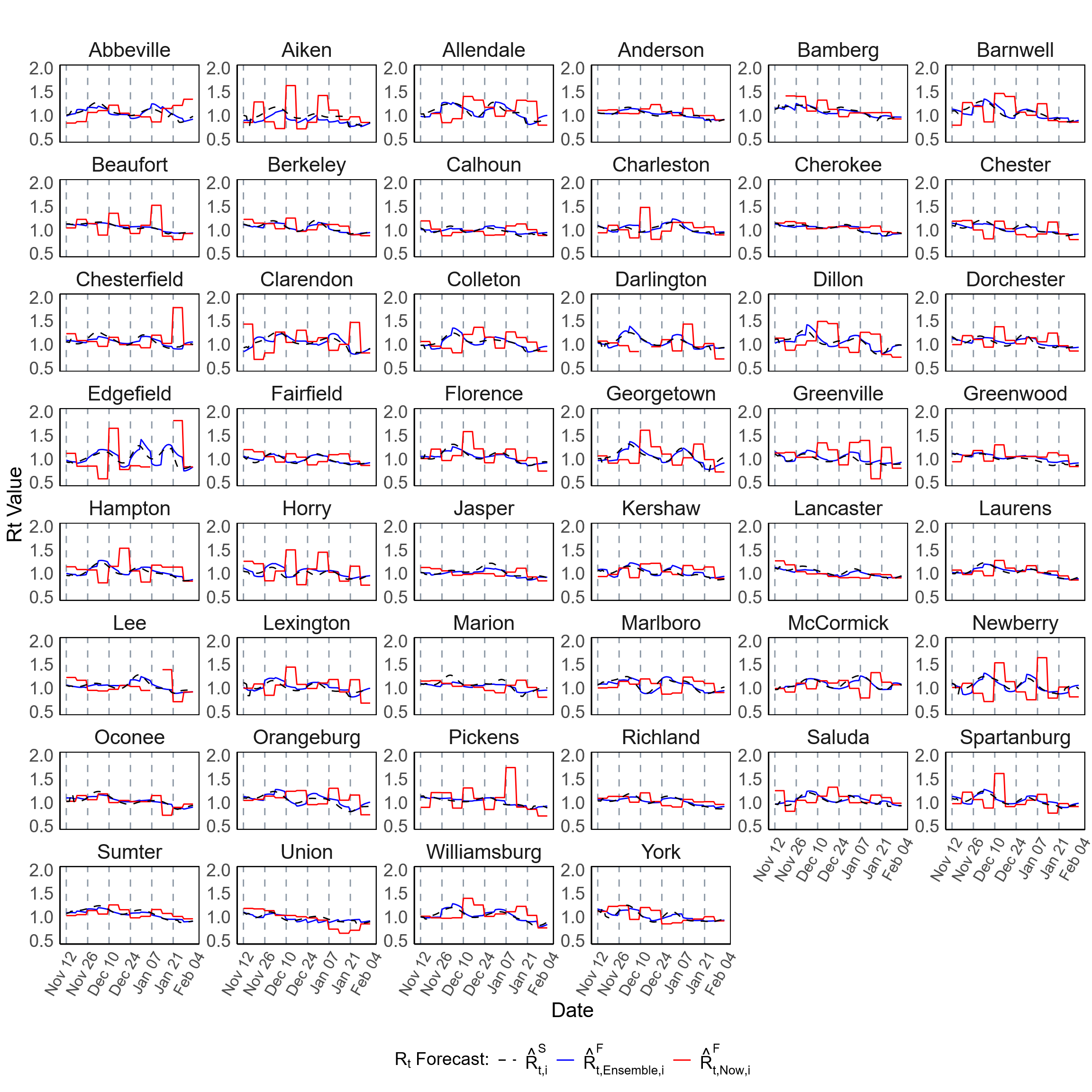

**Figure S1.** Forecast of $R_{t}$ at the county level in SC during Scenario-1 (November 11, 2020 – February 02, 2021). This figure presents the forecast of $R_{t}$ for all 46 counties in SC. The plots compare the ensemble-based forecast (blue lines) and forecasts generated using the EpiNow2 R package (red lines) alongside the spatially (covariate-adjusted) smoothed estimates ($\hat{R}_{t, Now,i}^{S}$, black dashed lines), where $i$ represents the county, $t$ denotes the time point (day), and “Now” refers to the EpiNow2 method. The forecasts were generated for 7-day horizon predictions over 84 days period using a rolling window approach.

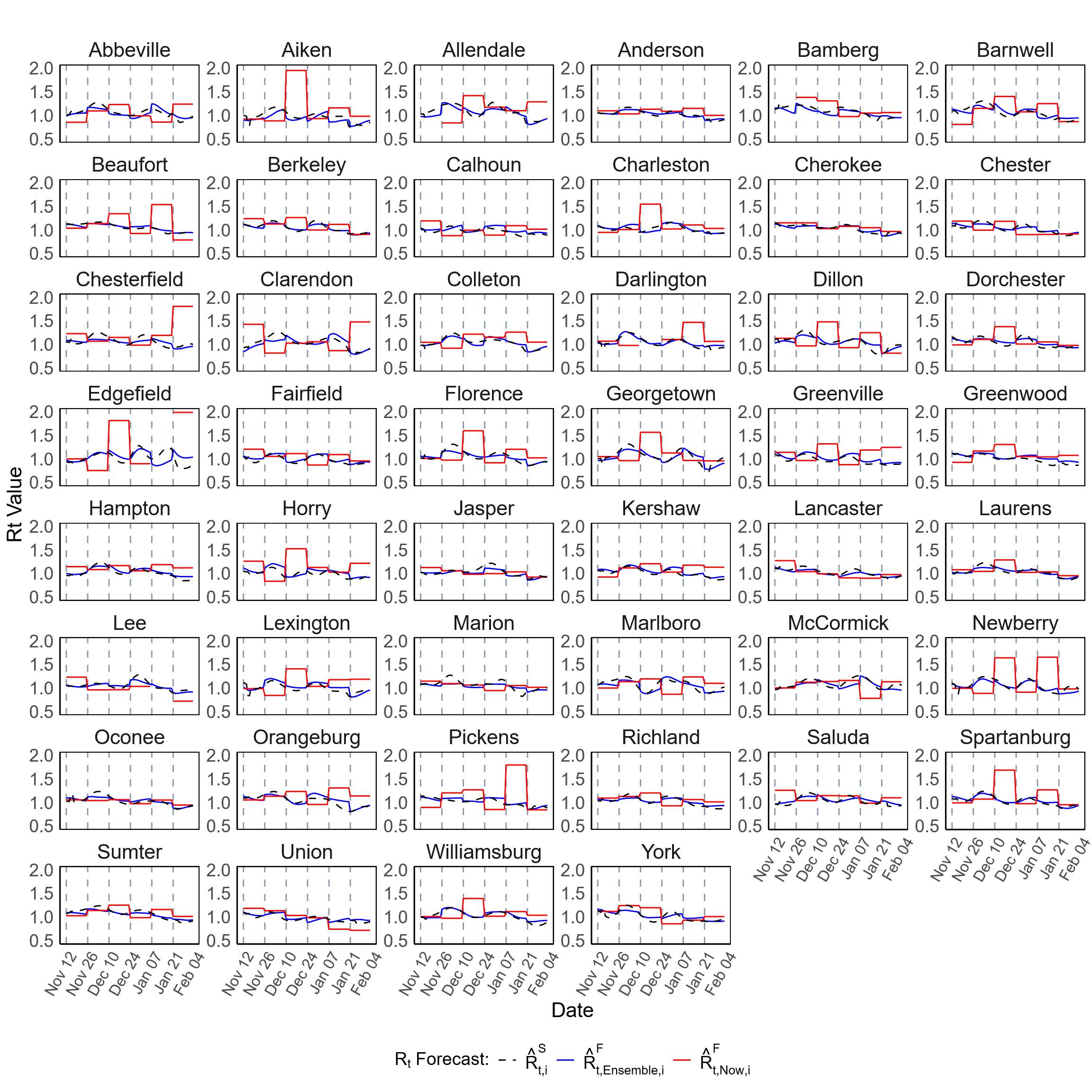

**Figure S2.** Forecast of $R_{t}$ at the county level in SC during Scenario-1 (November 11, 2020 – February 02, 2021). This figure presents the forecast of $R_{t}$ for all 46 counties in SC. The plots compare the ensemble-based forecast (blue lines) and forecasts generated using the EpiNow2 R package (red lines) alongside the spatially (covariate-adjusted) smoothed estimates ($\hat{R}_{t, Now,i}^{S}$, black dashed lines), where $i$ represents the county, $t$ denotes the time point (day), and “Now” refers to the EpiNow2 method. The forecasts were generated for 14-day horizon predictions over 84 days period using a rolling window approach.

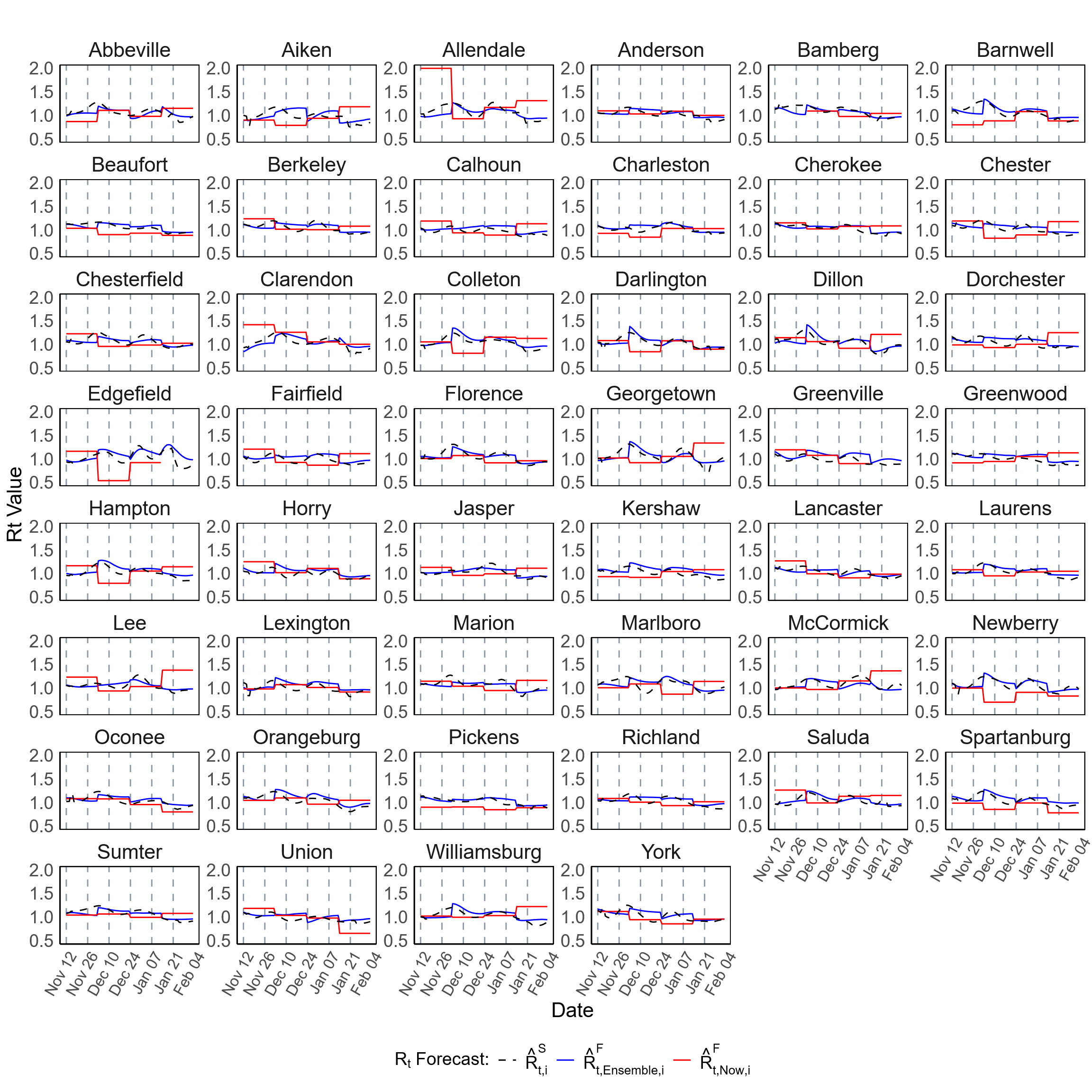

**Figure S3.** Forecast of $R_{t}$ at the county level in SC during Scenario-1 (November 11, 2020 – February 02, 2021). This figure presents the forecast of $R_{t}$ for all 46 counties in SC. The plots compare the ensemble-based forecast (blue lines) and forecasts generated using the EpiNow2 R package (red lines) alongside the spatially (covariate-adjusted) smoothed estimates ($\hat{R}_{t, Now,i}^{S}$, black dashed lines), where $i$ represents the county, $t$ denotes the time point (day), and “Now” refers to the EpiNow2 method. The forecasts were generated for 21-day horizon predictions over 84 days period using a rolling window approach.

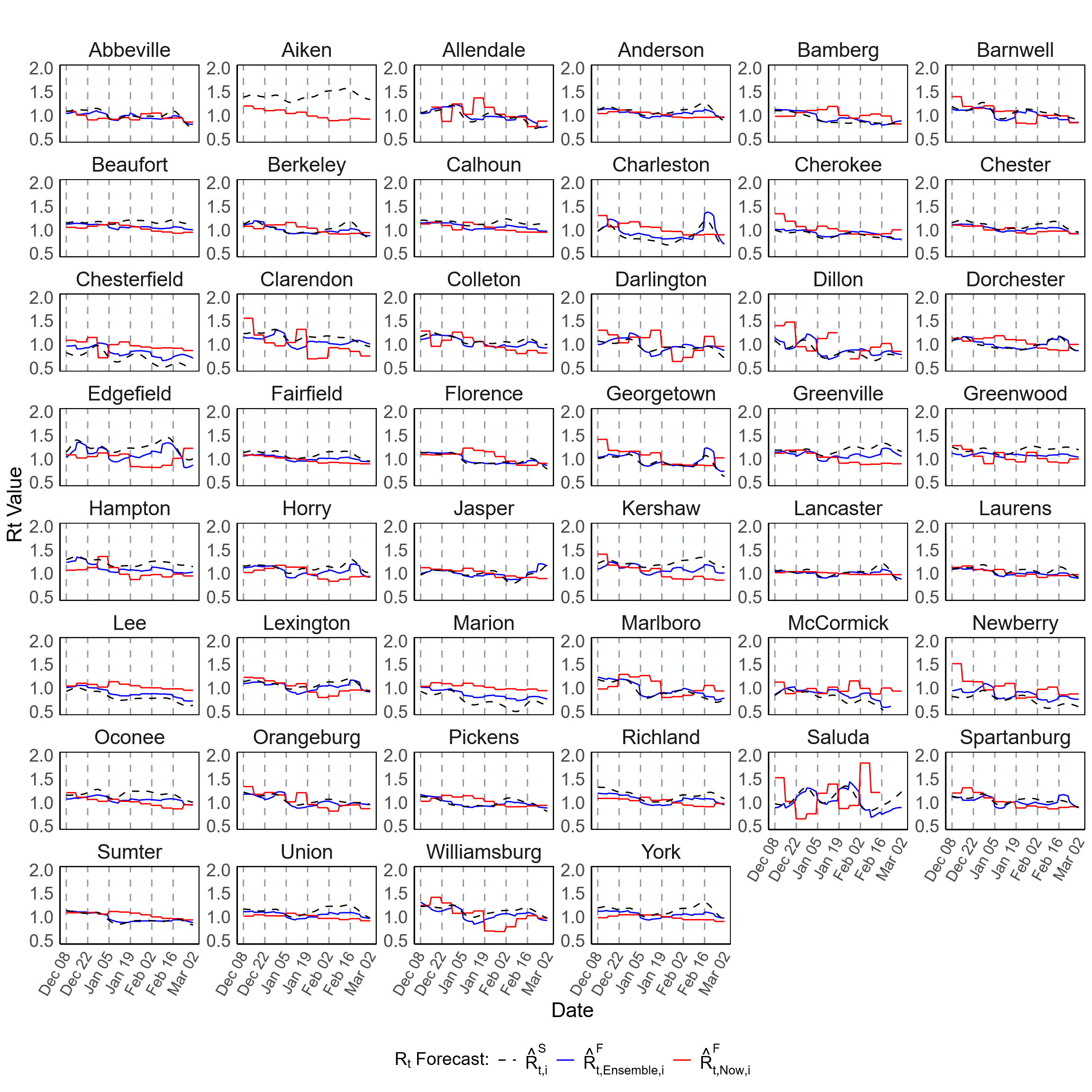

**Figure S4.** Forecast of $R_{t}$ at the county level in SC during Scenario-2 (December 11, 2022 – March 04, 2023). This figure presents the forecast of $R_{t}$ for all 46 counties in SC. The plots compare the ensemble-based forecast (blue lines) and forecasts generated using the EpiNow2 R package (red lines) alongside the spatially (covariate-adjusted) smoothed estimates ($\hat{R}_{t, Now,i}^{S}$, black dashed lines), where $i$ represents the county, $t$ denotes the time point (day), and “Now” refers to the EpiNow2 method. The forecasts were generated for 7-day horizon predictions over 84 days period using a rolling window approach.

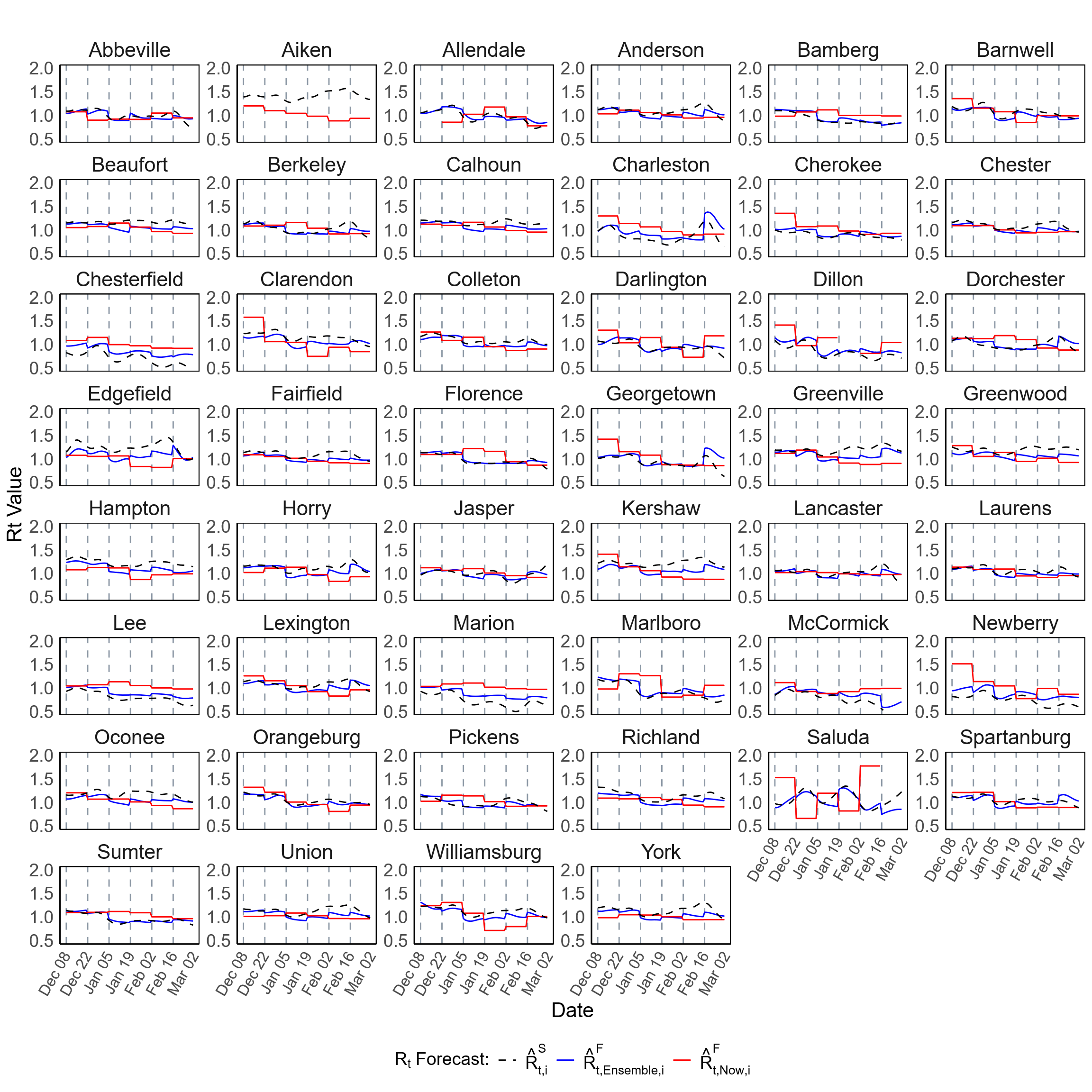

**Figure S5.** Forecast of $R_{t}$ at the county level in SC during Scenario-2 (December 11, 2022 – March 04, 2023). This figure presents the forecast of $R_{t}$ for all 46 counties in SC. The plots compare the ensemble-based forecast (blue lines) and forecasts generated using the EpiNow2 R package (red lines) alongside the spatially (covariate-adjusted) smoothed estimates ($\hat{R}_{t, Now,i}^{S}$, black dashed lines), where $i$ represents the county, $t$ denotes the time point (day), and “Now” refers to the EpiNow2 method. The forecasts were generated for 14-day horizon predictions over 84 days period using a rolling window approach.

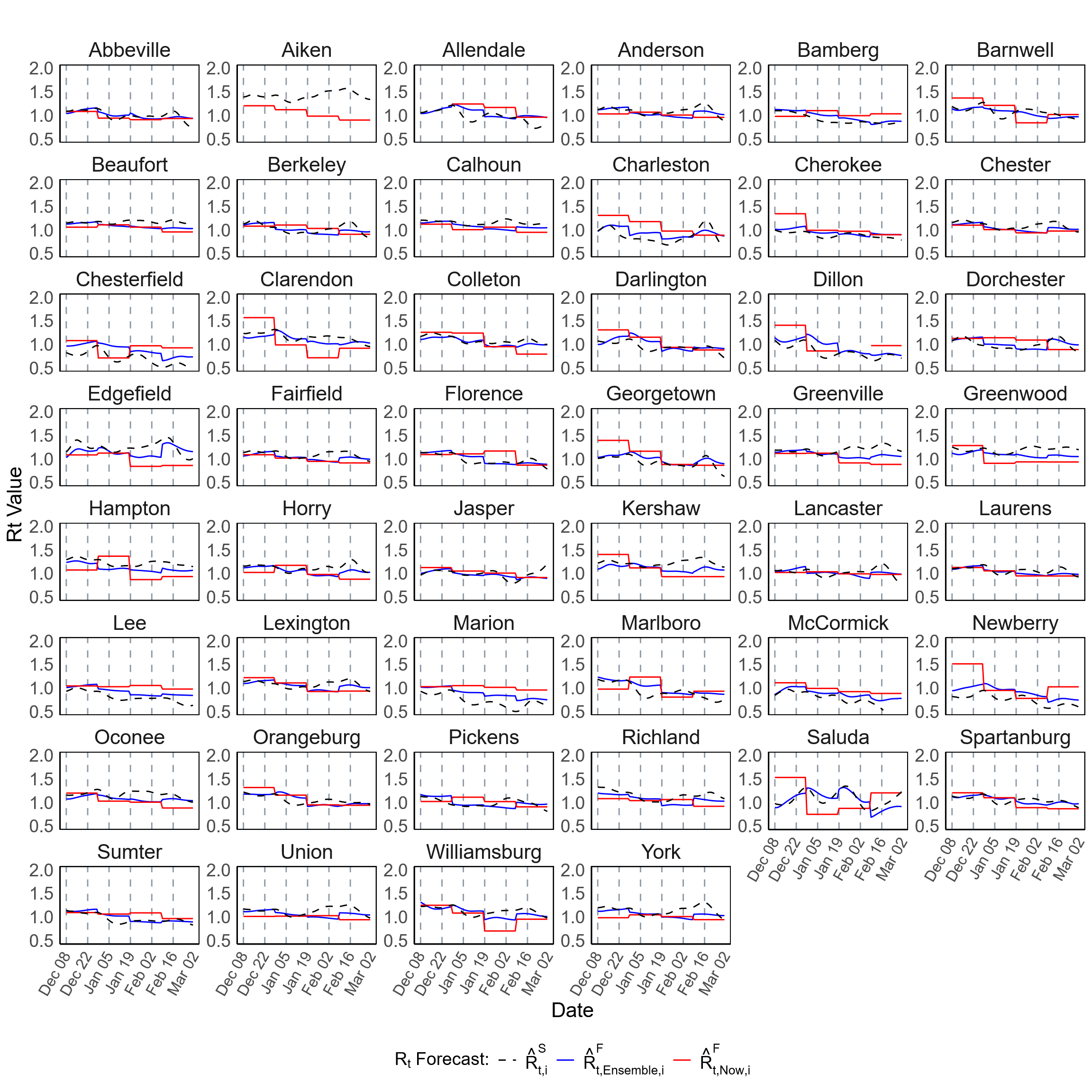

**Figure S6.** Forecast of $R_{t}$ at the county level in SC during Scenario-2 (December 11, 2022 – March 04, 2023). This figure presents the forecast of $R_{t}$ for all 46 counties in SC. The plots compare the ensemble-based forecast (blue lines) and forecasts generated using the EpiNow2 R package (red lines) alongside the spatially (covariate-adjusted) smoothed estimates ($\hat{R}_{t, Now,i}^{S}$, black dashed lines), where $i$ represents the county, $t$ denotes the time point (day), and “Now” refers to the EpiNow2 method. The forecasts were generated for 21-day horizon predictions over 84 days period using a rolling window approach.

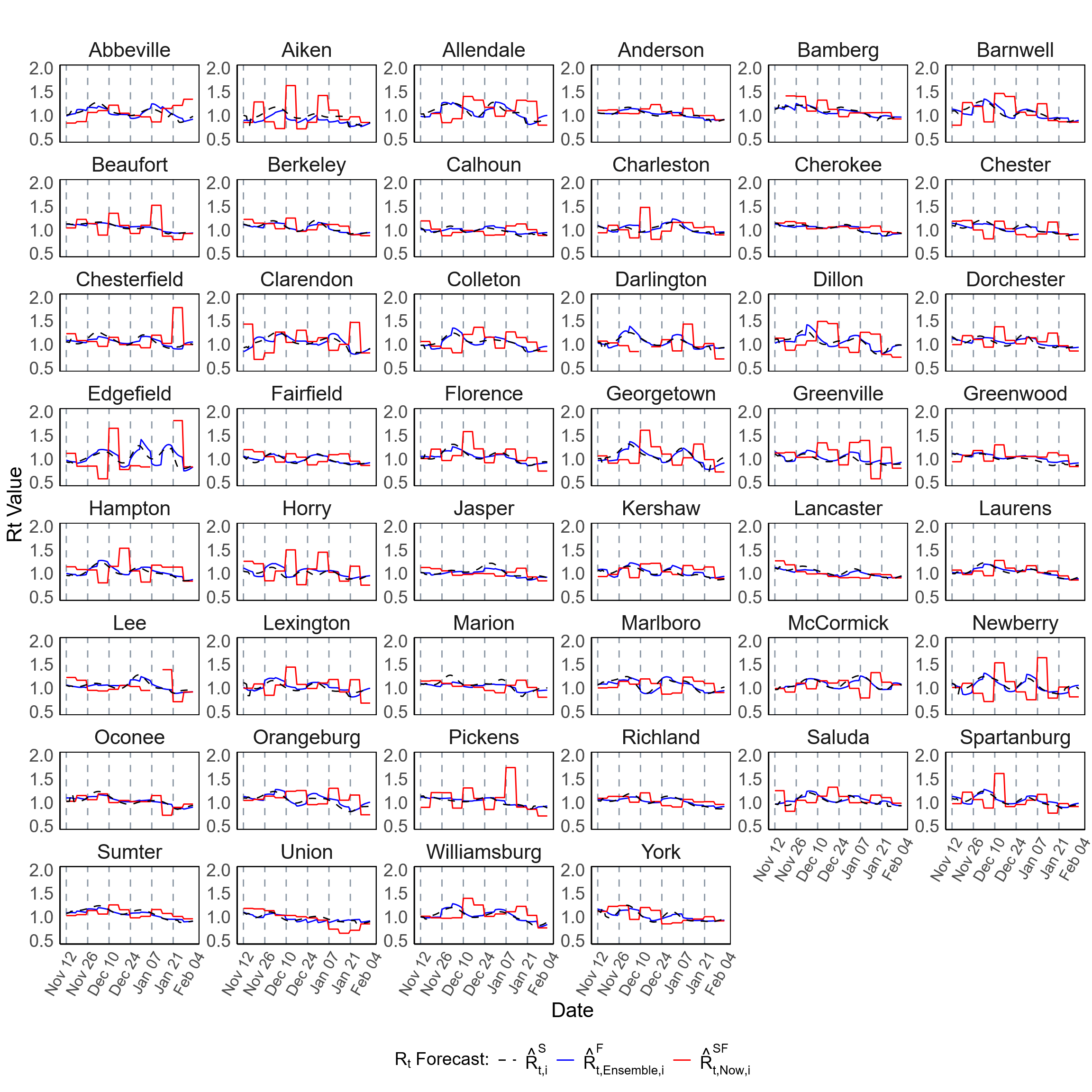

**Figure S7.** Forecast of $R_{t}$ at the county level in SC during Scenario-1 (November 11, 2020 – February 02, 2021). This figure presents the forecast of $R_{t}$ for all 46 counties in SC. The plots compare the ensemble-based forecast (blue lines) and forecasts generated using the EpiNow2 R package and spatially (covariate-adjusted) smoothed ($\hat{R}_{t, Now,i}^{\mathrm{SF}}$, red lines) alongside the spatially (covariate-adjusted) smoothed estimates ($\hat{R}_{t,i}^{S}$, black dashed lines), where $i$ represents the county, $t$ denotes the time point (day), and “Now” refers to the EpiNow2 method. The forecasts were generated for 7-day horizon predictions over 84 days period using a rolling window approach.

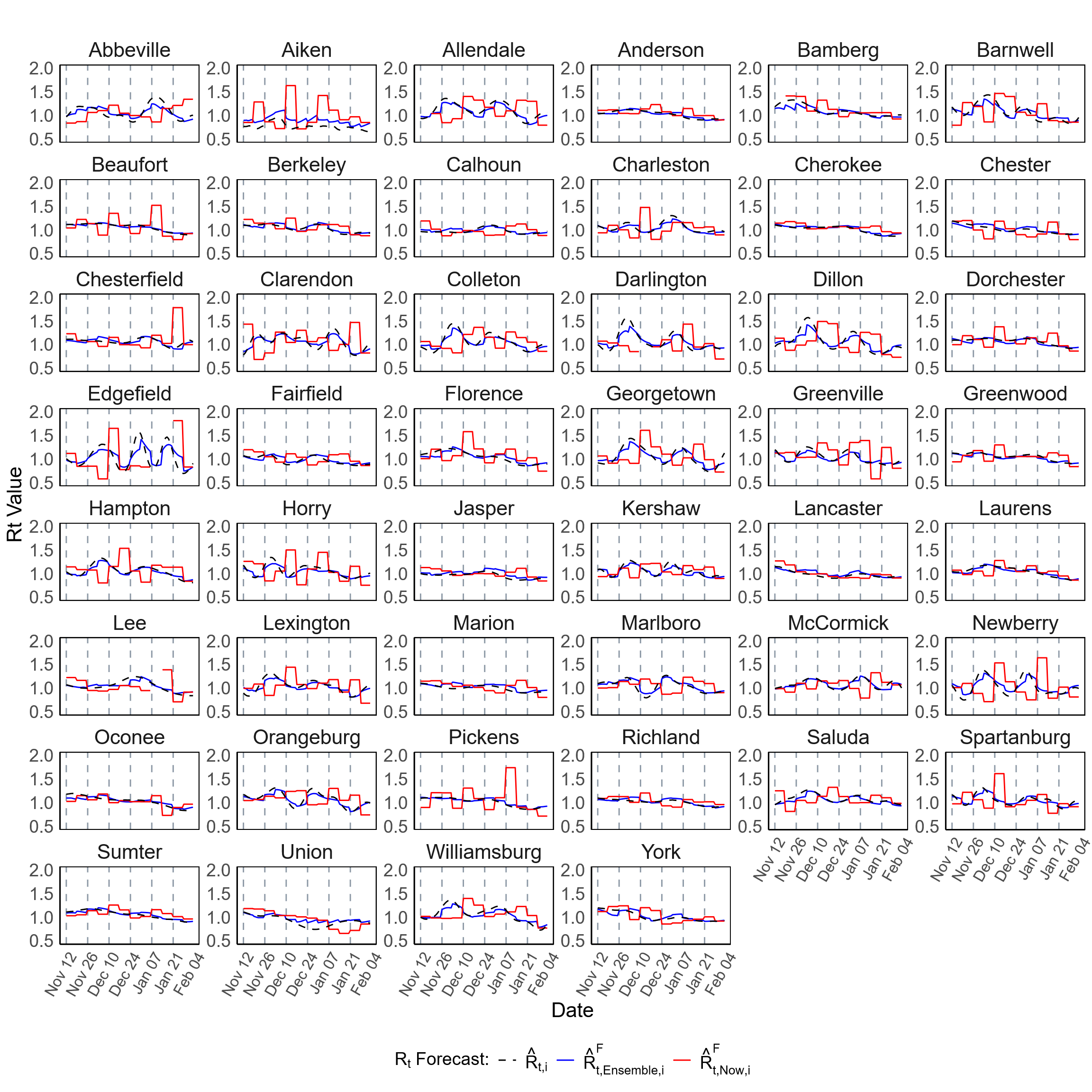

**Figure S8.** Forecast of $R_{t}$ at the county level in SC during Scenario-1 (November 11, 2020 – February 02, 2021). This figure presents the forecast of $R_{t}$ for all 46 counties in SC. The plots compare the ensemble-based forecast (blue lines), and forecasts generated using the EpiNow2 R package (red lines) alongside the initial estimates ( $\hat{R}_{t,i}$, black dashed lines), where $i$ represents the county, $t$ denotes the time point (day), and “Now” refers to the EpiNow2 method. The forecasts were generated for 7-day horizon predictions over 84 days period using a rolling window approach.

**Table S2*.*** Forecast accuracy metrics for the effective reproductive number $(R_{t})$ in SC counties during two forecast periods: November 11, 2020 – February 02, 2021 (Scenario-1) and December 11, 2022 – March 04, 2023 (Scenario-2). Here forecast accuracy was calculated comparing the $R_{t}$ forecast with the initial estimates of $R_{t}$. Forecast models were trained with data from June 01, 2020, to November 10, 2020, for Scenario-1, and from May 01, 2022, to December 10, 2022, for Scenario-2. The forecasting approach employed a rolling window method for 7-day, 14-day, and 21-day horizon predictions over 84 days period. The table presents the median PA across counties along with the IQR of county PAs.

| Forecast Method | Effective Reproductive Number Forecast:  Percentage Agreement (PA), Median (IQR) | | | | | |
| --- | --- | --- | --- | --- | --- | --- |
|  | Forecast Period: November 11, 2020 – February 02, 2021 (Scenario-1) | | | Forecast Period: December 11, 2022 – March 04, 2023 (Scenario-2) | | |
|  | 7-day horizon | 14-day horizon | 21-day horizon | 7-day horizon | 14-day horizon | 21-day horizon |
| EpiNow2 | 85.7% (82.0% – 90.8%) | 84.6% (80.4% – 90.2%) | 87.2% (82.9% – 90.8%) | 88.6% (86.9% – 90.5%) | 88.5% (83.2% – 91.6%) | 88.3% (83.2% – 92.1%) |
| Ensemble | 96.2% (95.0% – 97.1%) | 95.3% (93.0% – 96.4%) | 90.8% (89.7% – 92.4%) | 92.2% (90.1% – 94.3%) | 91.3% (89.5% – 92.8%) | 90.0% (88.0% – 91.8%) |

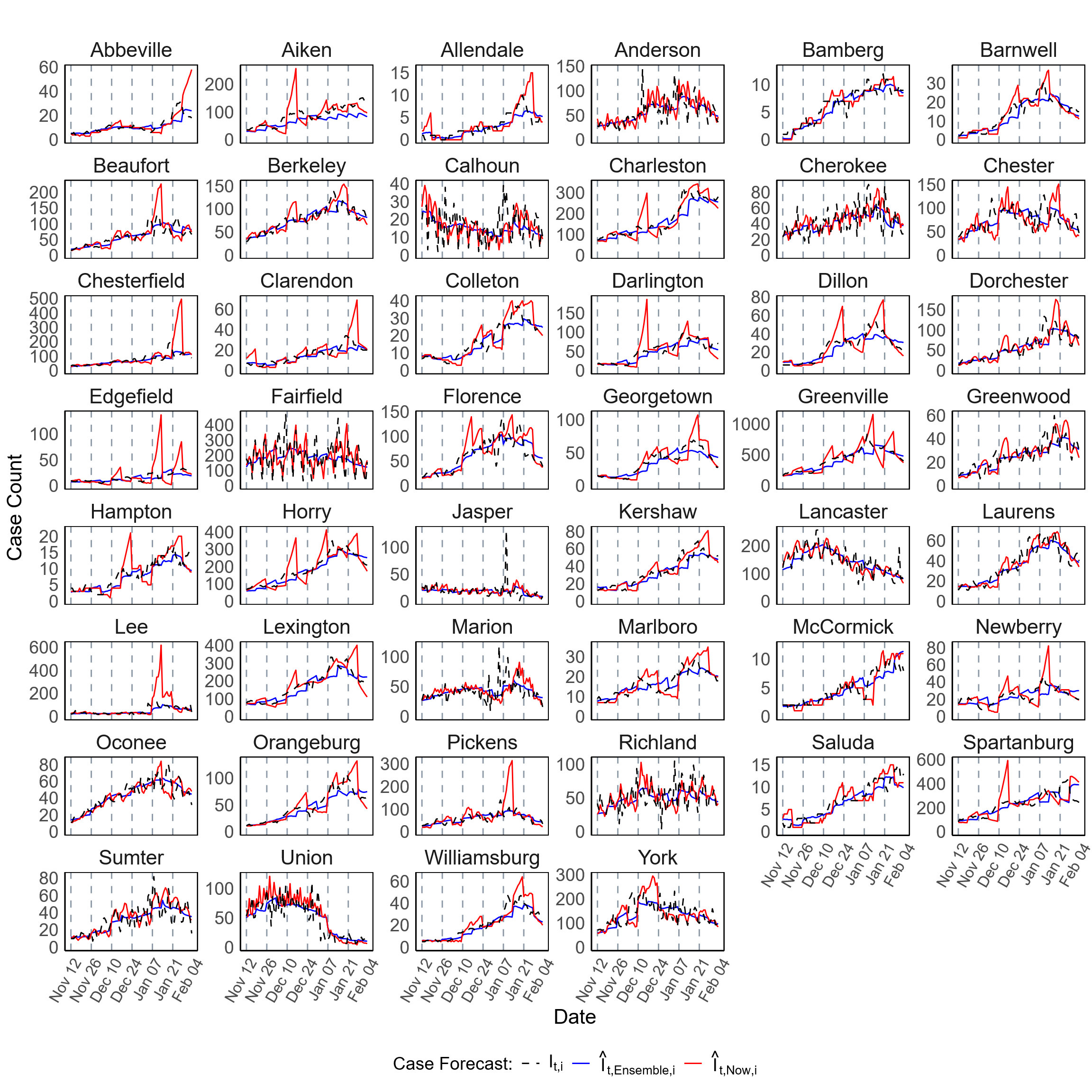

**Figure S9.** Forecast of COVID-19 case counts at the county level in SC during Scenario-1 (November 11, 2020 – February 02, 2021). This figure presents the forecast for COVID-19 case counts for all 46 counties in SC. The plots compare EpiNow2 forecasts ($\hat{I}_{t,Now,i}$, red lines) and ensemble-based forecasts ($\hat{I}_{t,Ensemble,i}$, blue lines) against the observed daily case counts ($I_{t,i}$,black dashed lines). The forecasts were generated for 7-day horizon predictions over 84 days period using a rolling window approach.

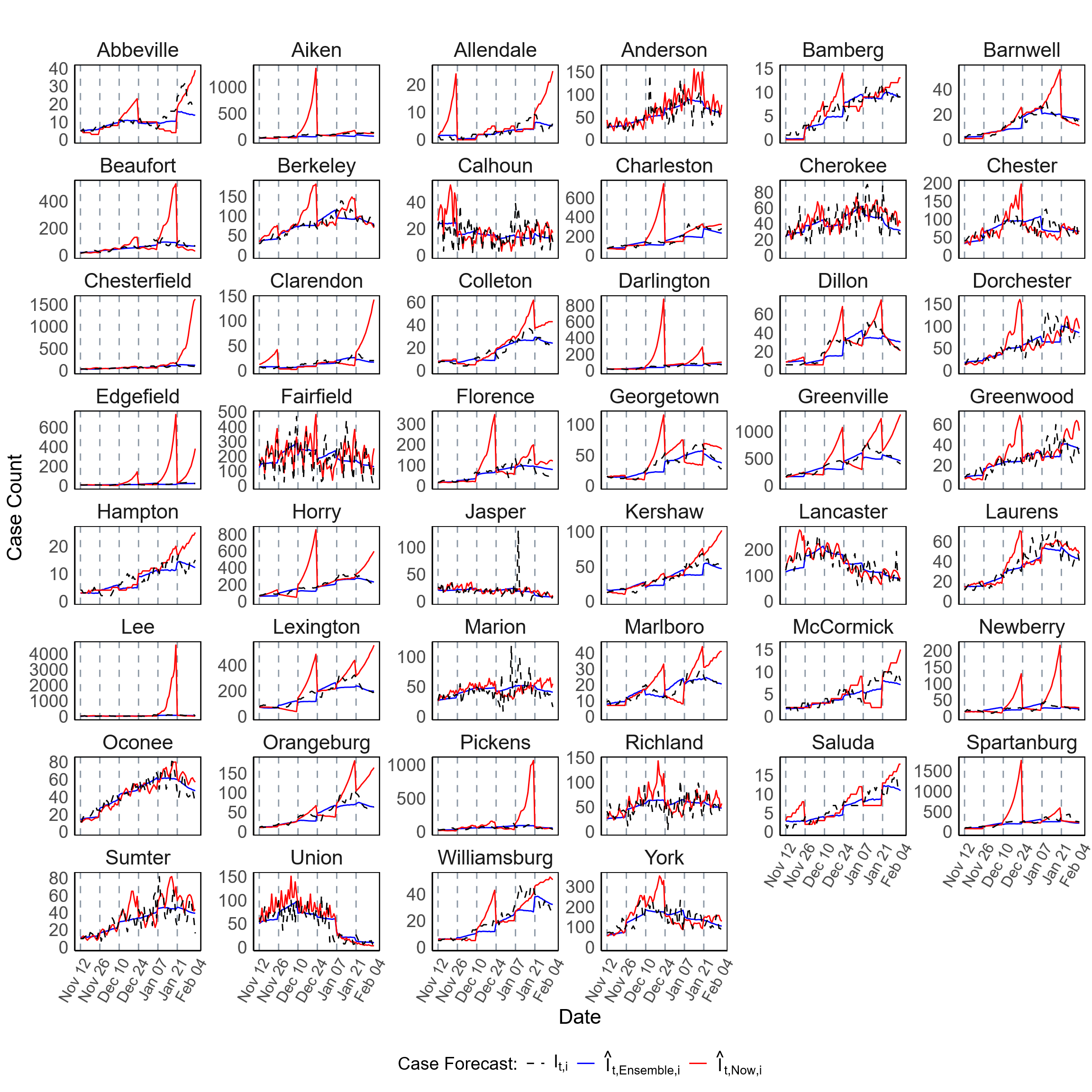

**Figure S10.** Forecast of COVID-19 case counts at the county level in SC during Scenario-1 (November 11, 2020 – February 02, 2021). This figure presents the forecast for COVID-19 case counts for all 46 counties in SC. The plots compare EpiNow2 forecasts ($\hat{I}_{t,Now,i}$, red lines) and ensemble-based forecasts ($\hat{I}_{t,Ensemble,i}$, blue lines) against the observed daily case counts ($I_{t,i}$,black dashed lines). The forecasts were generated for 14-day horizon predictions over 84 days period using a rolling window approach.

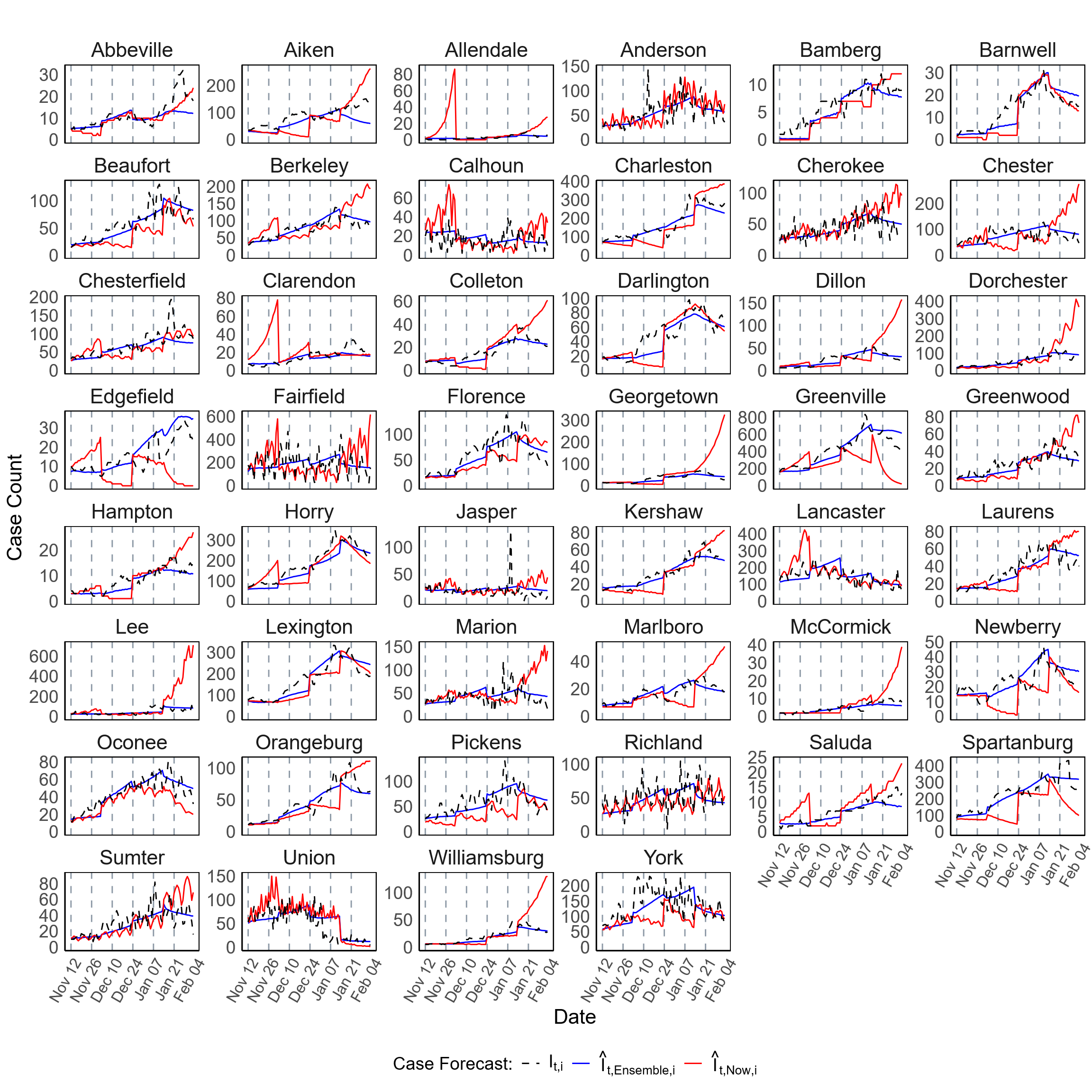

**Figure S11.** Forecast of COVID-19 case counts at the county level in SC during Scenario-1 (November 11, 2020 – February 02, 2021). This figure presents the forecast for COVID-19 case counts for all 46 counties in SC. The plots compare EpiNow2 forecasts ($\hat{I}_{t,Now,i}$, red lines) and ensemble-based forecasts ($\hat{I}_{t,Ensemble,i}$, blue lines) against the observed daily case counts ($I_{t,i}$,black dashed lines). The forecasts were generated for 21-day horizon predictions over 84 days period using a rolling window approach.

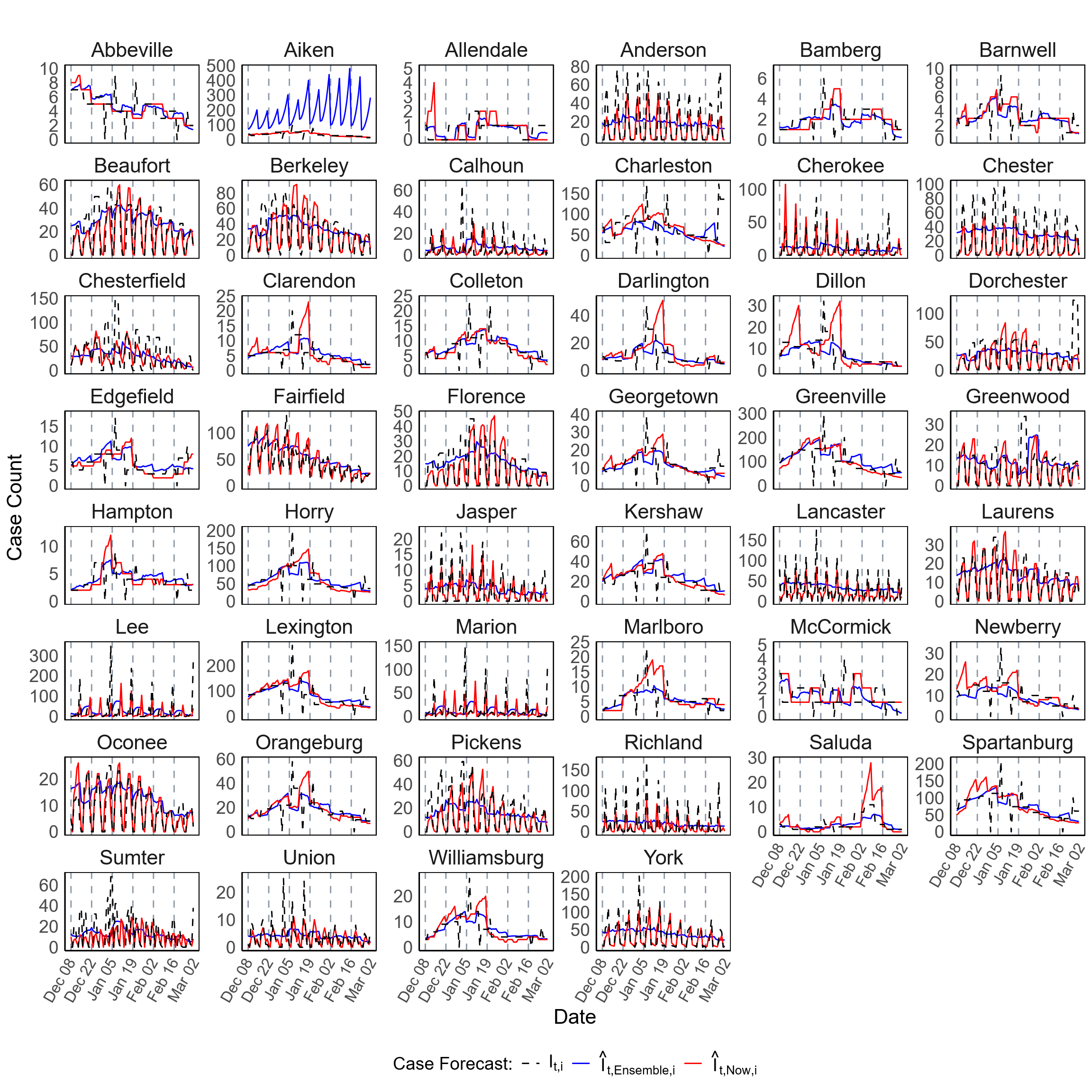

**Figure S12.** Forecast of COVID-19 case counts at the county level in SC during Scenario-2 (December 11, 2022 – March 04, 2023). This figure presents the forecast for COVID-19 case counts for all 46 counties in SC. The plots compare EpiNow2 forecasts ($\hat{I}_{t,Now,i}$, red lines) and ensemble-based forecasts ($\hat{I}_{t,Ensemble,i}$, blue lines) against the observed daily case counts ($I_{t,i}$,black dashed lines). The forecasts were generated for 7-day horizon predictions over 84 days period using a rolling window approach.

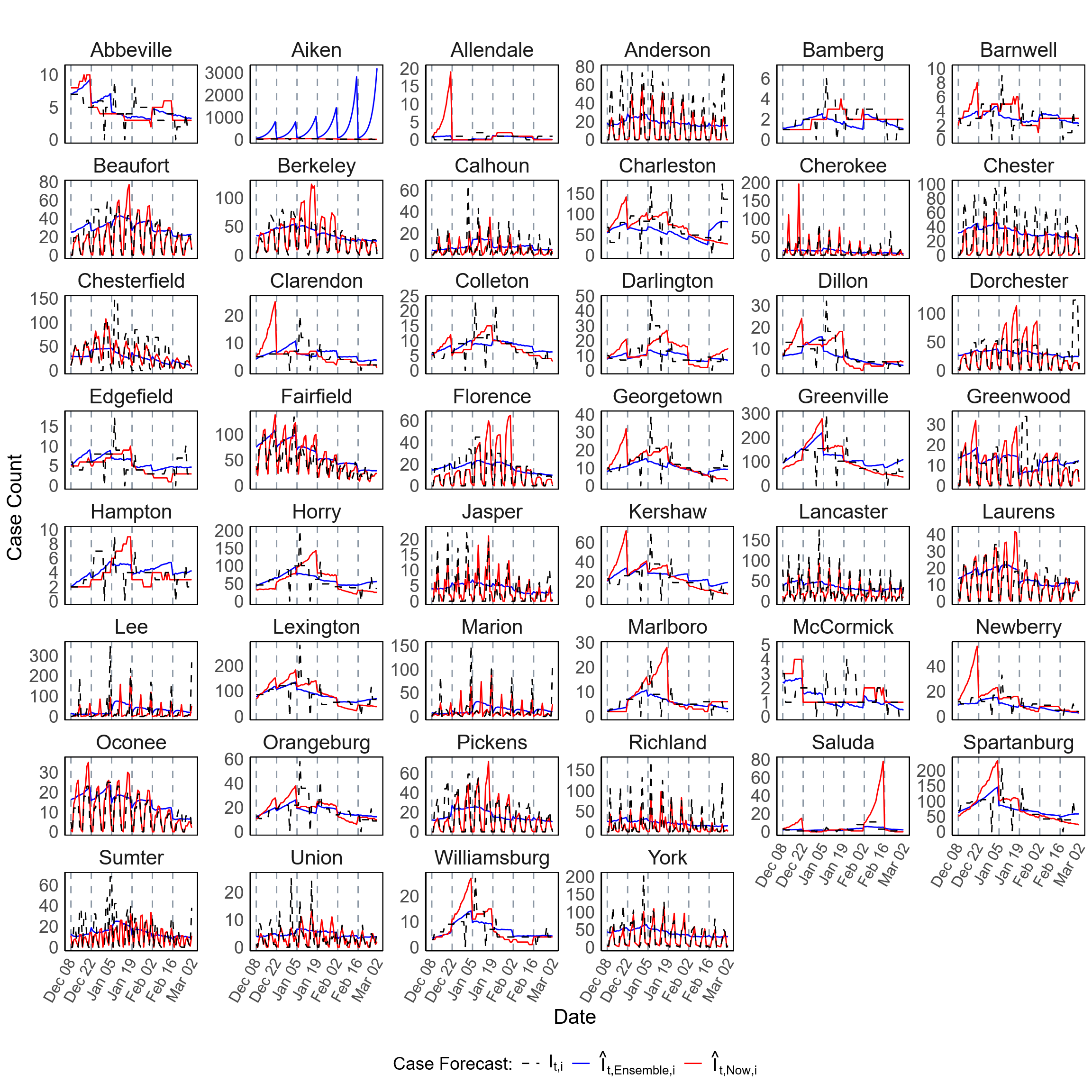

**Figure S13.** Forecast of COVID-19 case counts at the county level in SC during Scenario-2 (December 11, 2022 – March 04, 2023). This figure presents the forecast for COVID-19 case counts for all 46 counties in SC. The plots compare EpiNow2 forecasts ($\hat{I}_{t,Now,i}$, red lines) and ensemble-based forecasts ($\hat{I}_{t,Ensemble,i}$, blue lines) against the observed daily case counts ($I_{t,i}$,black dashed lines). The forecasts were generated for 14-day horizon predictions over 84 days period using a rolling window approach.

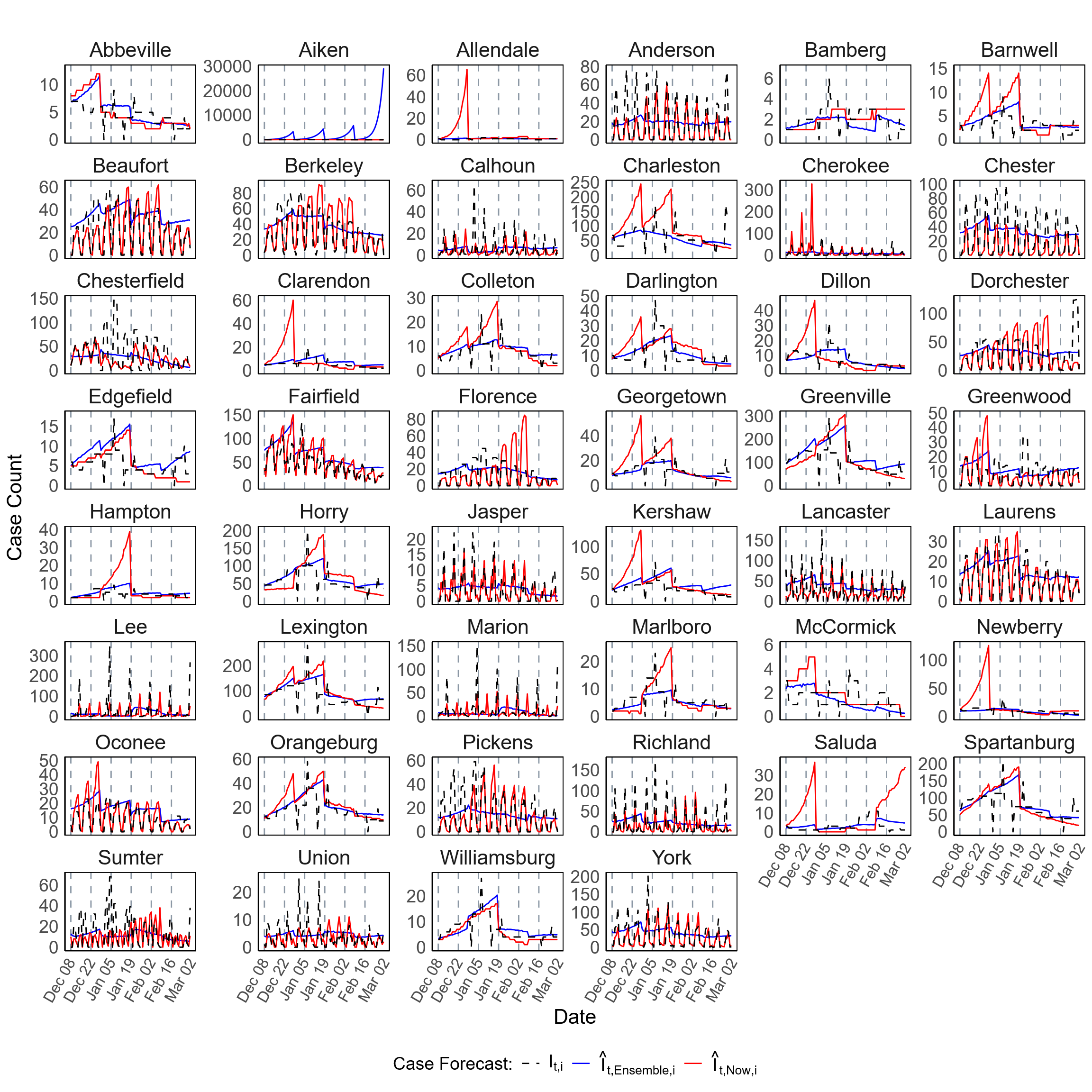

**Figure S14.** Forecast of COVID-19 case counts at the county level in SC during Scenario-2 (December 11, 2022 – March 04, 2023). This figure presents the forecast for COVID-19 case counts for all 46 counties in SC. The plots compare EpiNow2 forecasts ($\hat{I}_{t,Now,i}$, red lines) and ensemble-based forecasts ($\hat{I}_{t,Ensemble,i}$, blue lines) against the observed daily case counts ($I_{t,i}$,black dashed lines). The forecasts were generated for 21-day horizon predictions over 84 days period using a rolling window approach.

**Table S3.** Forecast accuracy metrics for individual models used in the ensemble-based forecasting of $R_{t}$ at the county level in SC during Scenario-1 (November 11, 2020 – February 02, 2021) and Scenario-2 (December 11, 2022 – March 04, 2023). The models include Reg, RF, and XGB, applied to both the initial estimates, $\hat{R}_{t,Now,i}$, and the spatially (covariate-adjusted) smoothed estimates, $\hat{R}_{t, Now,i}^{S}$. Forecast accuracy is evaluated using PA with median and IQR. The forecasting approach involved 7-day, 14-day, and 21-day horizon predictions over 84 days period using a rolling window method. The accuracy metrics for each individual forecasting model are summarized in this table.

| Forecast Method | Effective Reproductive Number Forecast:  Percentage Agreement (PA), Median (IQR) | | | | | |
| --- | --- | --- | --- | --- | --- | --- |
|  | Forecast Period: November 11, 2020 – February 02, 2021 | | | Forecast Period: December 11, 2022 – March 04, 2023 | | |
|  | 7-day horizon | 14-day horizon | 21-day horizon | 7-day horizon | 14-day horizon | 21-day horizon |
| Regression | 94.5% (93.5% – 95.9%) | 95.0% (94.2% – 95.8%) | 94.10% (92.6.6% – 94.9%) | 86.5% (82.6% – 90.8%) | 86.7% (82.7% – 90.7%) | 86.6% (83.4% – 89.9%) |
| Regression (Smooth) | 97.3% (96.7% – 97.7%) | 95.5% (94.6% – 96.6%) | 94.5% (93.8% – 95.0%) | 98.6% (98.3% – 98.8%) | 95.8% (95.1% – 96.5%) | 94.3% (93.3% – 95.3%) |
| RF | 94.1% (92.2% – 95.2%) | 94.2% (92.4% – 95.3%) | 93.1% (91.8% – 94.9%) | 86.7% (82.4% – 91.5%) | 88.8% (82.5% – 91.2%) | 87.4% (84.0% – 90.5%) |
| RF (Smooth) | 96.4% (95.6% – 96.7%) | 95.4% (94.2% – 96.1%) | 92.8% (91.8% – 93.3%) | 96.6% (95.3% – 97.2%) | 94.7% (93.3% – 95.8%) | 93.2% (91.7% – 94.5%) |
| XGBoost | 94.2% (92.9% – 95.6%) | 94.0% (93.0% – 95.0%) | 92.8% (91.5% – 94.1%) | 86.5% (83.4% – 91.0%) | 86.5% (83.2% – 91.4%) | 87.5% (83.9% – 90.5%) |
| XGBoost (Smooth) | 97.3% (96.8% – 97.7%) | 95.9% (94.8% – 96.3%) | 93.4% (92.4% – 94.2%) | 97.8% (97.2% – 98.2%) | 94.7% (93.9% – 96.2%) | 91.7% (90.2% – 93.5%) |
| EpiNow2 (Smooth) | 87.6% (86.6% – 88.9%) | 86.2% (83.8% – 88.2%) | 89.7% (87.5% – 92.3%) | 87.2% (84.0% – 89.9%) | 86.7% (84.2% – 89.9%) | 87.5% (84.8% – 90.3%) |

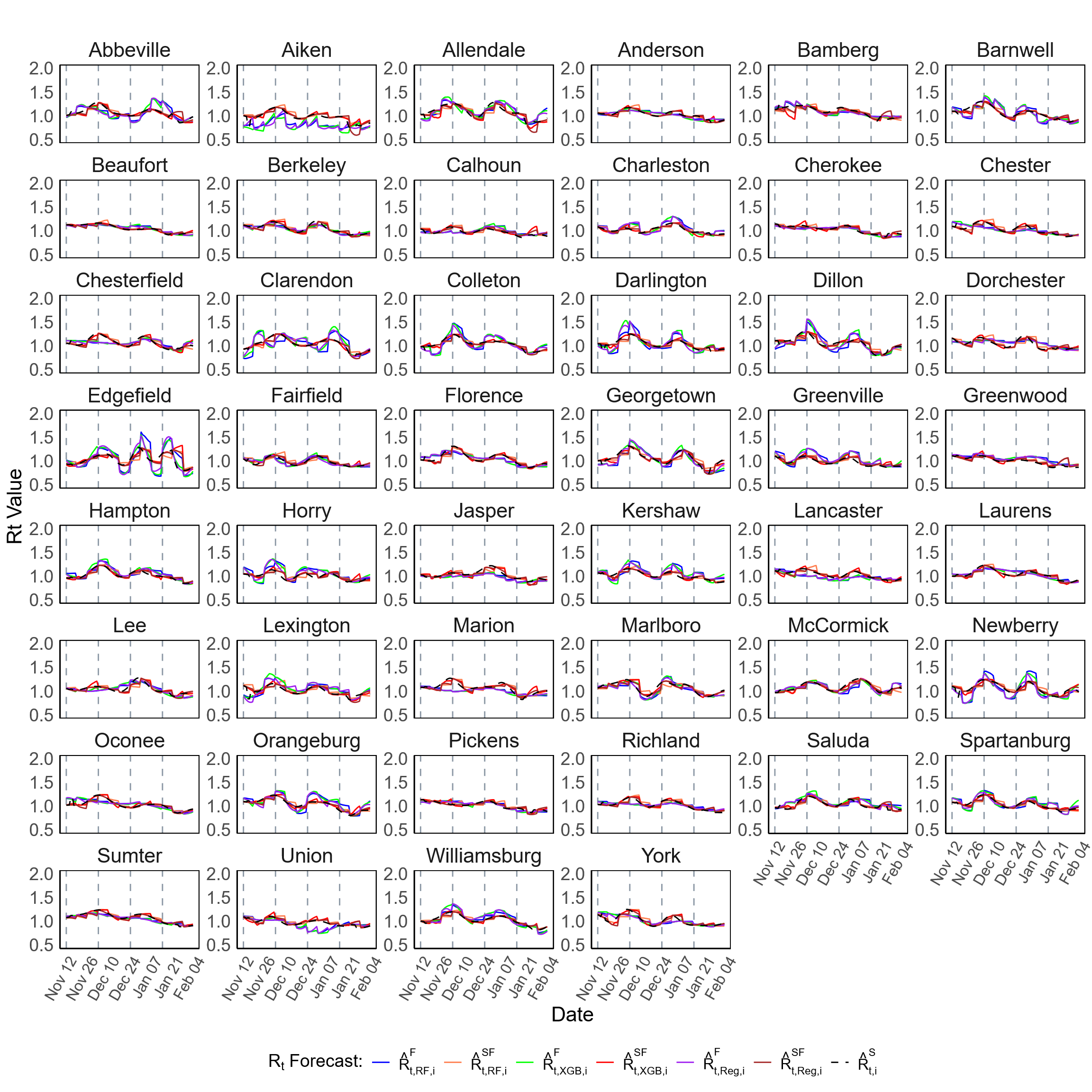

**Figure S15.** Forecast of the effective reproductive number $(R_{t})$ at the county level in SC during Scenario-1 (November 11, 2020 – February 02, 2021). This figure presents the forecast of $R_{t}$ for all 46 counties in SC. The plots compare forecasts generated using multiple individual models, alongside the spatially (covariate-adjusted) smoothed estimates (black dashed lines,  $\hat{R}_{t,i}^{S}$), where $i$presents the county, $t$ denotes the time point (day), and “Now” refers to the **EpiNow2** method. The forecasts were generated for 7-day horizon predictions over 84 days period using a rolling window approach.

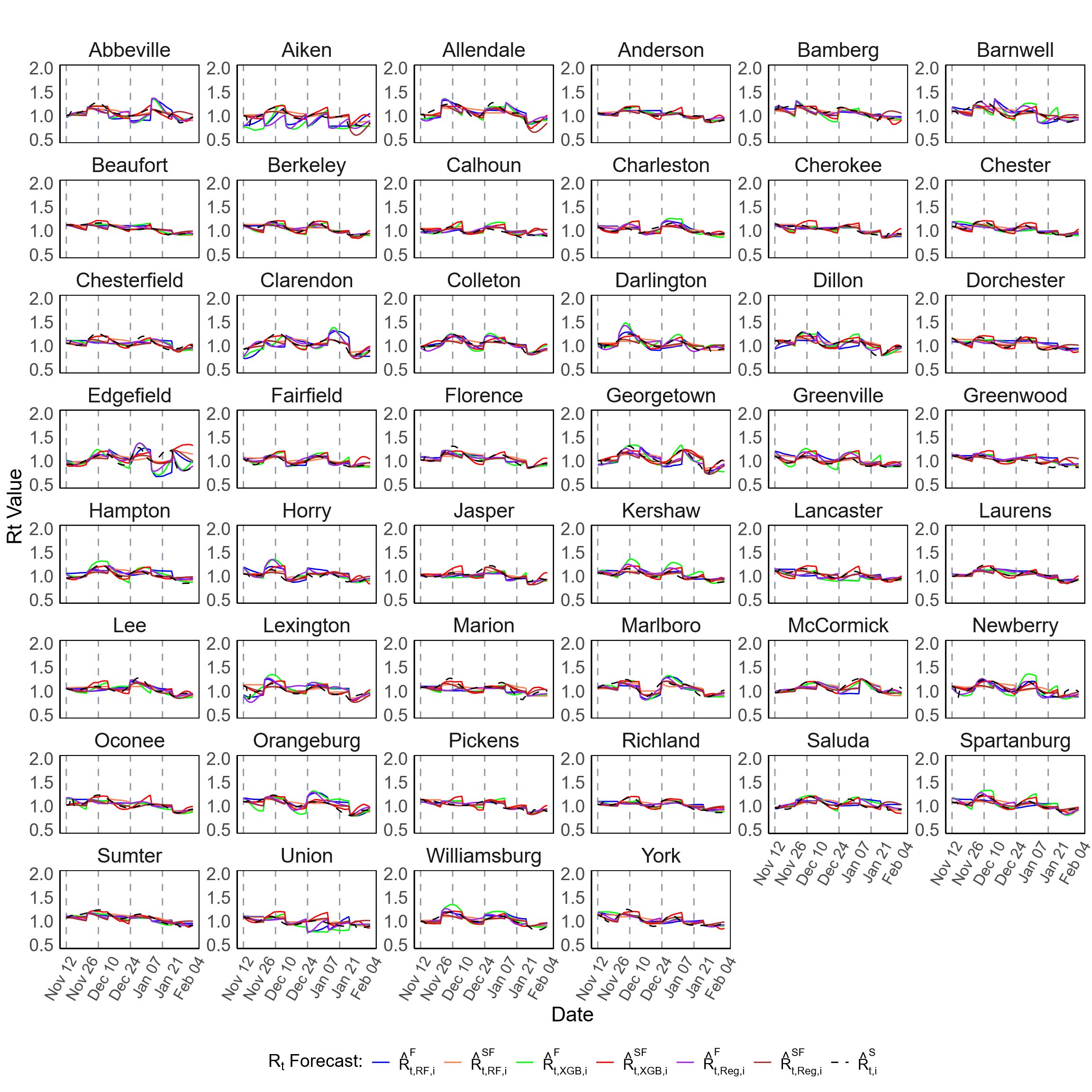

**Figure S16.** Forecast of the effective reproductive number $(R_{t})$ at the county level in SC during Scenario-1 (November 11, 2020 – February 02, 2021). This figure presents the forecast of $R_{t}$ for all 46 counties in SC. The plots compare forecasts generated using multiple individual models, alongside the spatially (covariate-adjusted) smoothed estimates (black dashed lines,  $\hat{R}_{t,i}^{S}$), where $i$presents the county, $t$ denotes the time point (day), and “Now” refers to the **EpiNow2** method. The forecasts were generated for 14-day horizon predictions over 84 days period using a rolling window approach.

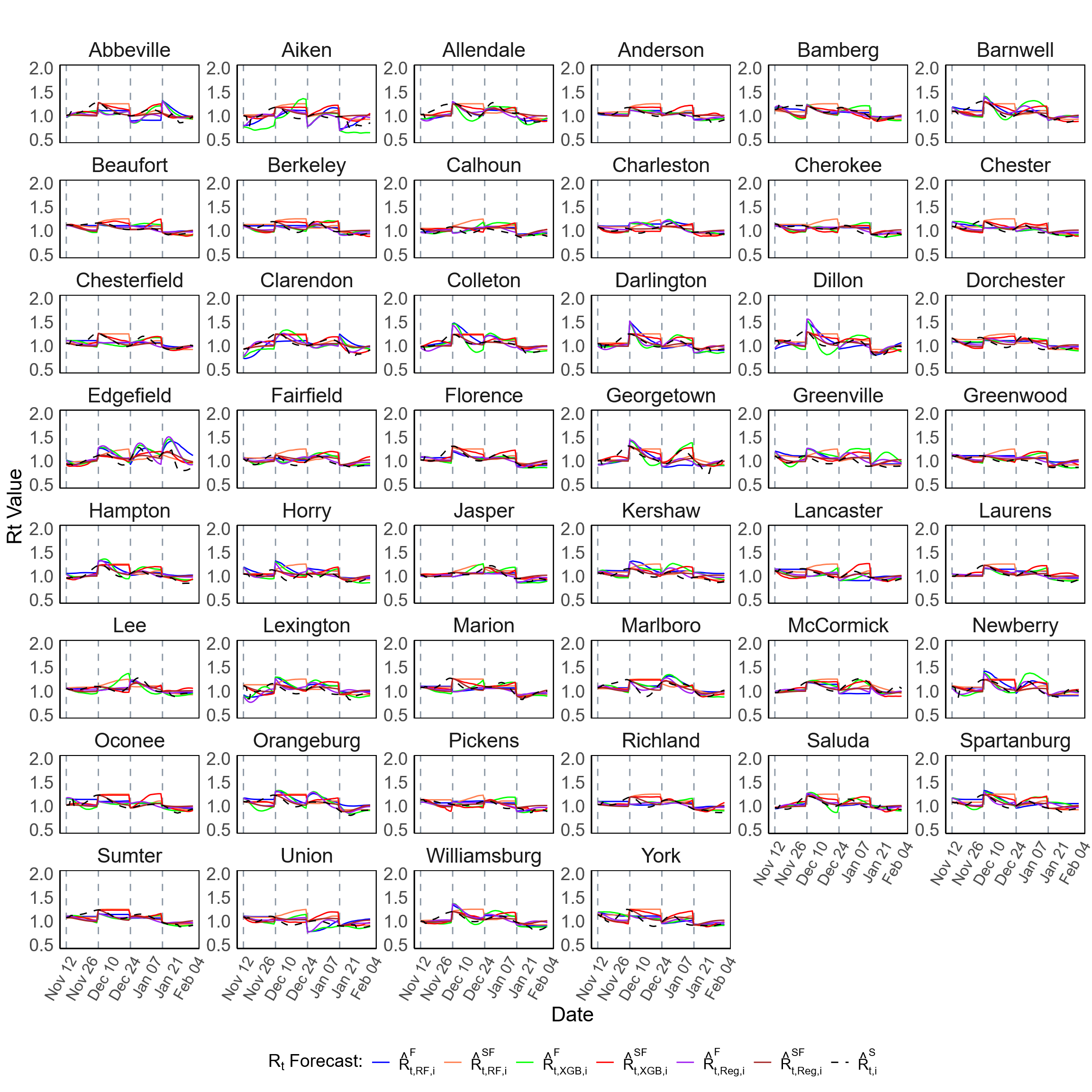

**Figure S17.** Forecast of the effective reproductive number $(R_{t})$ at the county level in SC during Scenario-1 (November 11, 2020 – February 02, 2021). This figure presents the forecast of $R_{t}$ for all 46 counties in SC. The plots compare forecasts generated using multiple individual models, alongside the spatially (covariate-adjusted) smoothed estimates (black dashed lines,  $\hat{R}_{t,i}^{S}$), where $i$presents the county, $t$ denotes the time point (day), and “Now” refers to the **EpiNow2** method. The forecasts were generated for 21-day horizon predictions over 84 days period using a rolling window approach.

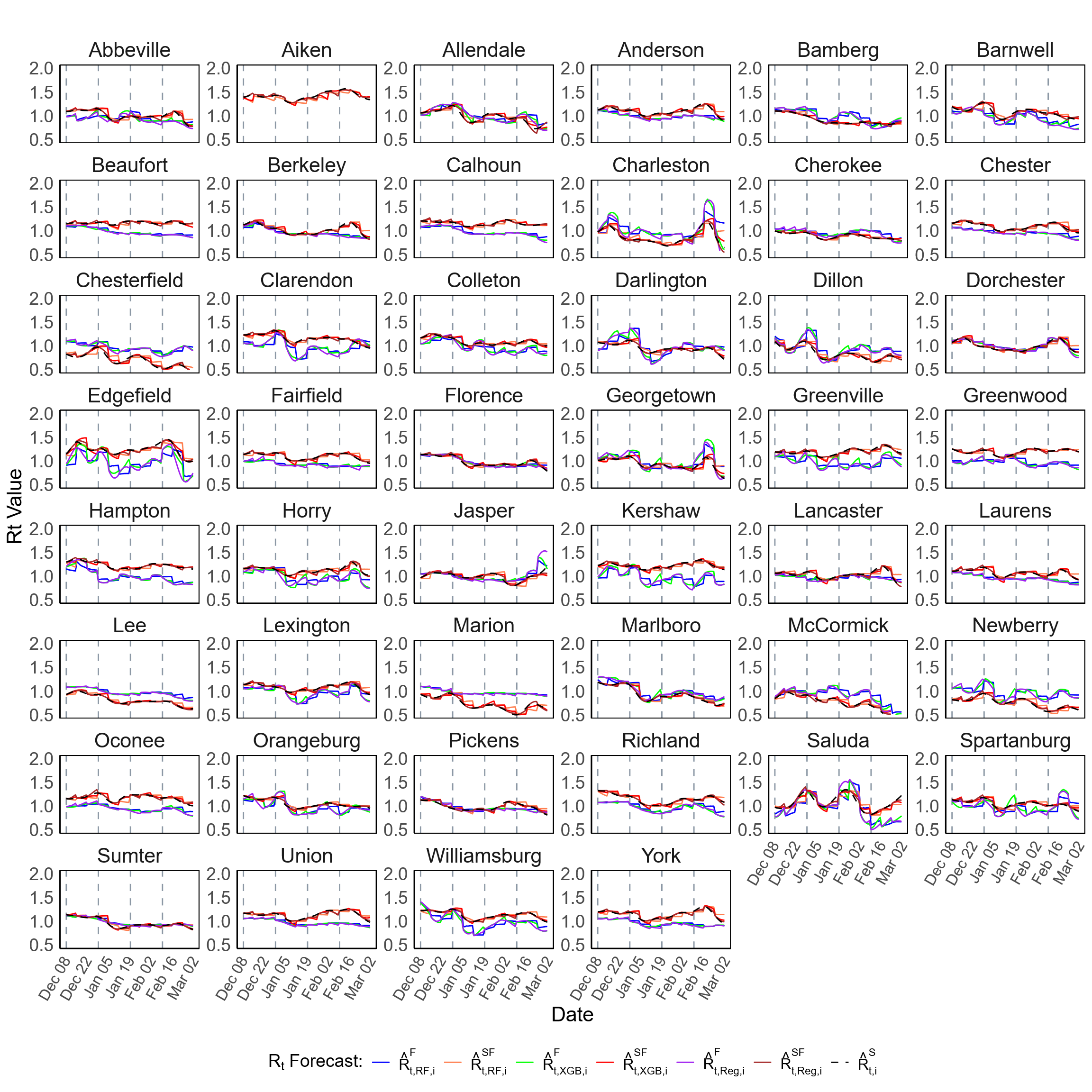

**Figure S18.** Forecast of the effective reproductive number $(R_{t})$ at the county level in SC during Scenario-2 (December 11, 2022 – March 04, 2023). This figure presents the forecast of $R_{t}$ for all 46 counties in SC. The plots compare forecasts generated using multiple individual models, alongside the spatially (covariate-adjusted) smoothed estimates (black dashed lines,  $\hat{R}_{t,i}^{S}$), where $i$presents the county, $t$ denotes the time point (day), and “Now” refers to the **EpiNow2** method. The forecasts were generated for 7-day horizon predictions over 84 days period using a rolling window approach.

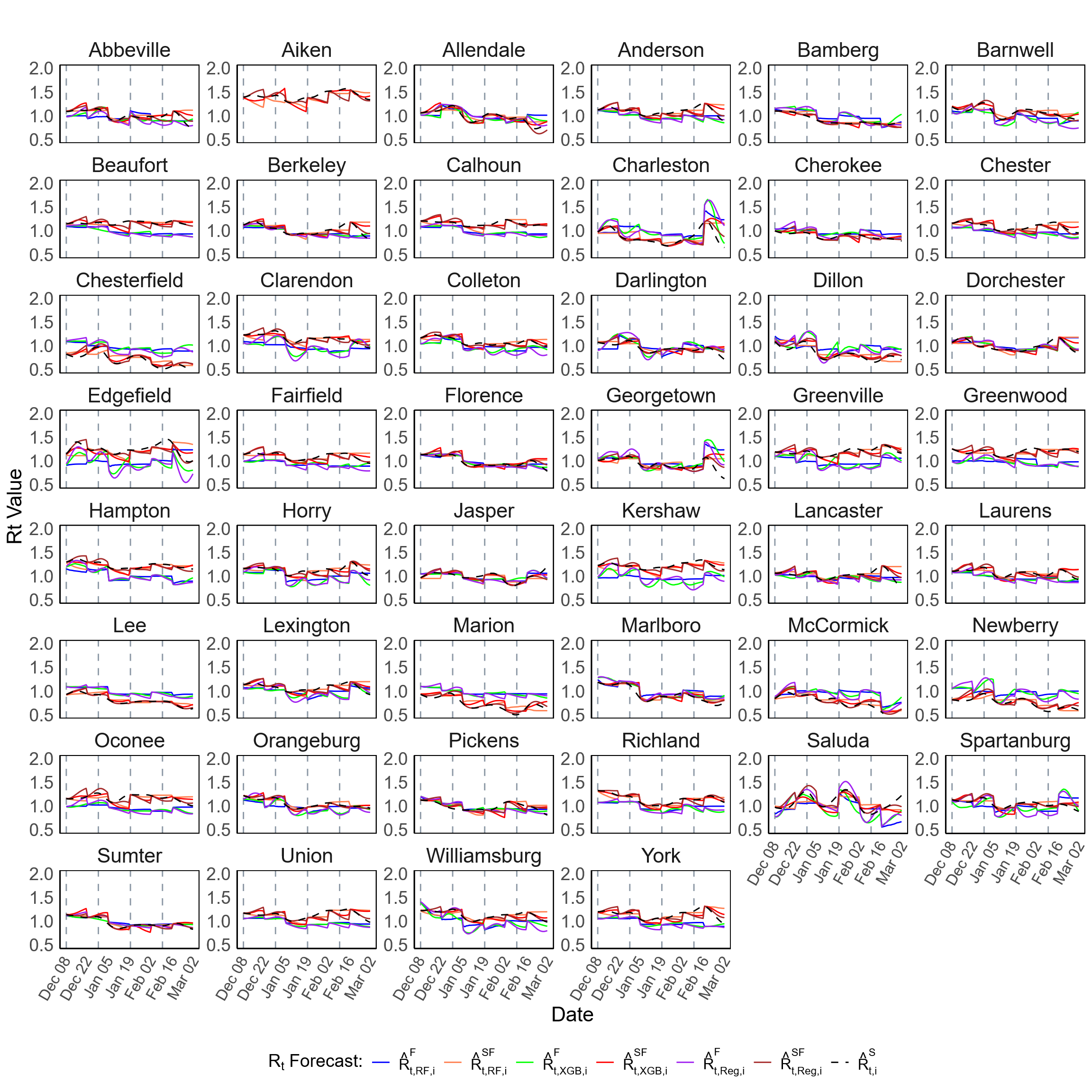

**Figure S19.** Forecast of the effective reproductive number $(R_{t})$ at the county level in SC during Scenario-2 (December 11, 2022 – March 04, 2023). This figure presents the forecast of $R_{t}$ for all 46 counties in SC. The plots compare forecasts generated using multiple individual models, alongside the spatially (covariate-adjusted) smoothed estimates (black dashed lines,  $\hat{R}_{t,i}^{S}$), where $i$presents the county, $t$ denotes the time point (day), and “Now” refers to the **EpiNow2** method. The forecasts were generated for 14-day horizon predictions over 84 days period using a rolling window approach.

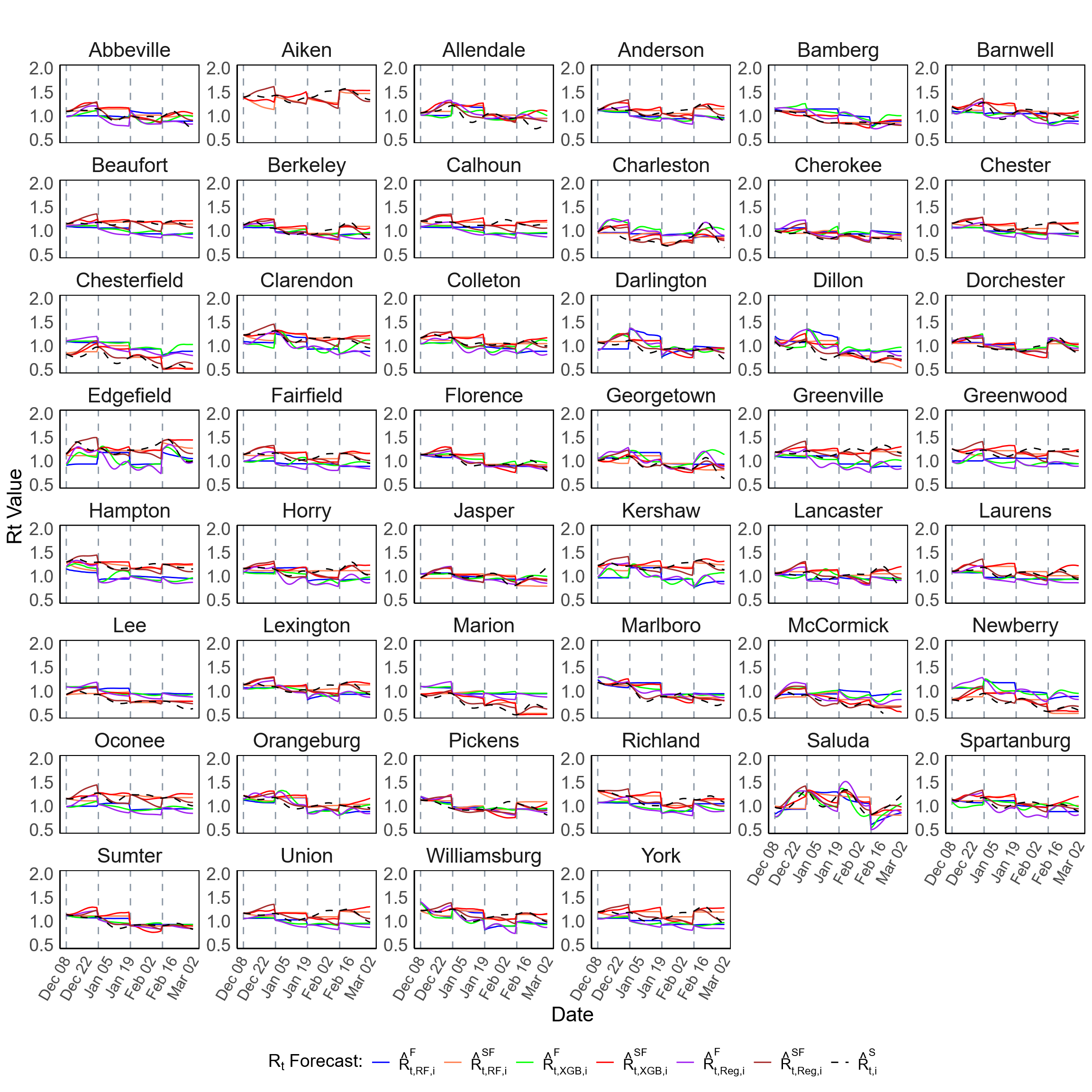

**Figure S20.** Forecast of the effective reproductive number $(R_{t})$ at the county level in SC during Scenario-2 (December 11, 2022 – March 04, 2023). This figure presents the forecast of $R_{t}$ for all 46 counties in SC. The plots compare forecasts generated using multiple individual models, alongside the spatially (covariate-adjusted) smoothed estimates (black dashed lines,  $\hat{R}_{t,i}^{S}$), where $i$presents the county, $t$ denotes the time point (day), and “Now” refers to the **EpiNow2** method. The forecasts were generated for 21-day horizon predictions over 84 days period using a rolling window approach.

**Table S4.** Forecast accuracy metrics for COVID-19 daily case counts forecasts using individual models at the county level in SC during Scenario-1 (November 11, 2020 – February 02, 2021) and Scenario-2 (December 11, 2022 – March 04, 2023). The last row presents accuracy of $R_{t}$ forecasting using EpiNow2, followed by spatial (covariate-adjusted) smoothing. We employed a probabilistic approach using a Poisson model to generate case forecasts based on the forecasted $R_{t}$ values, including $\hat{R}_{t,Reg,i}^{F}$, $\hat{R}_{t,RF,i}^{F}$, $\hat{R}_{t,XGB,i}^{F}$, $\hat{R}_{t,Reg,i}^{SF}$, $\hat{R}_{t,RF,i}^{SF}$, and $\hat{R}_{t,XGB,i}^{SF}$, then evaluated by comparing with observed case counts. Accuracy was measured using PA with median and IQR. The forecasts were generated for 7-day, 14-day, and 21-day horizon predictions over 84 days period using a rolling window approach.

| Forecast Method | COVID-19 Daily Cases Forecast:  Percentage Agreement (PA), Median (IQR) | | | | | |
| --- | --- | --- | --- | --- | --- | --- |
|  | Forecast Period: November 11, 2020 – February 02, 2021 | | | Forecast Period: December 11, 2022 – March 04, 2023 | | |
|  | 7-day horizon | 14-day horizon | 21-day horizon | 7-day horizon | 14-day horizon | 21-day horizon |
| Regression | 85.8% (83.2% – 88.2%) | 84.1% (81.3% – 86.8%) | 80.7% (78.6% – 84.7%) | 78.7% (73.1% – 82.6%) | 80.0% (74.6% – 82.7%) | 79.4% (75.2% – 82.7%) |
| Regression (Smooth) | 86.7% (83.1% – 87.8%) | 83.5% (80.8% – 86.5%) | 79.8% (78.4% – 82.9%) | 73.5% (67.9% – 78.0%) | 70.1% (62.6% – 76.2%) | 64.8% (56.6% – 73.0%) |
| RF | 87.1% (85.5% – 89.2%) | 85.2% (83.9% – 88.3%) | 84.1% (80.9% – 87.1%) | 81.4% (75.2% – 84.5%) | 82.4% (75.8% – 86.0%) | 79.5% (75.6% – 58.9%) |
| RF (Smooth) | 87.7% (84.9% – 89.7%) | 85.3% (82.6% – 87.4%) | 83.3% (78.4% – 86.2%) | 73.5% (69.8% – 78.9%) | 70.7% (64.3% – 76.4%) | 64.7% (58.9% – 73.8%) |
| XGBoost | 86.1% (83.3% – 88.3%) | 84.1% (81.4% – 87.9%) | 81.9% (77.9% – 85.1%) | 78.2% (72.7% – 81.3%) | 79.3% (74.7% – 83.1%) | 78.1% (73.4% – 81.9%) |
| XGBoost (Smooth) | 86.8% (83.4% – 87.8%) | 82.2% (79.9% – 85.0%) | 79.3% (77.5% – 83.6%) | 72.7% (68.2% – 77.9%) | 72.0% (63.0% – 76.5%) | 61.9% (54.8% – 72.3%) |

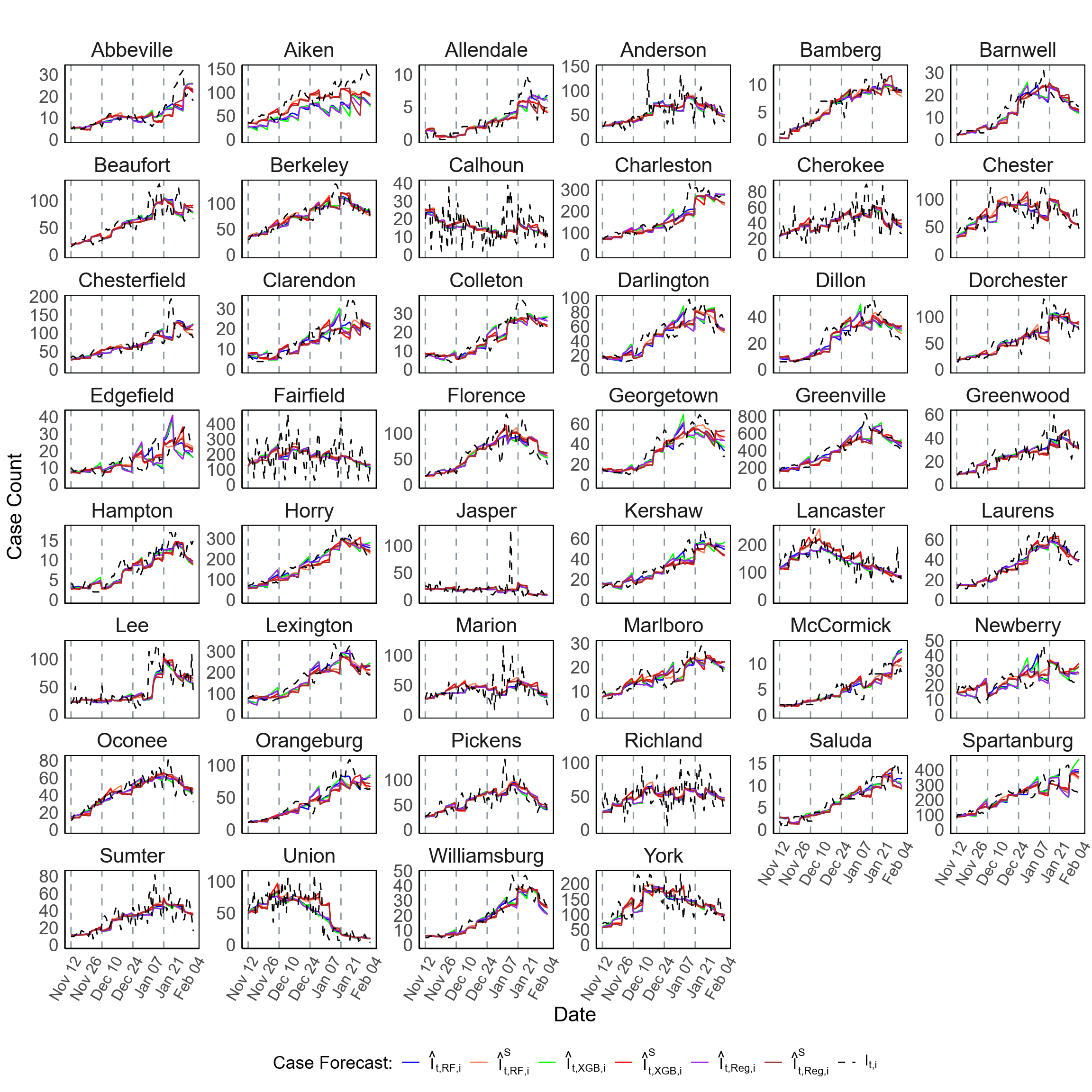

**Figure S21.** Forecast of COVID-19 case counts at the county level in SC during Scenario-1 (November 11, 2020 – February 02, 2021). This figure presents the forecast of COVID-19 case counts for all 46 counties in SC. The plots compare forecasts generated using multiple individual models against the observed daily case counts (black dashed lines, $I_{t,i}$), where $i$ represents the county and $t$ denotes the time point (day). The forecasts were generated for 7-day horizon predictions over 84 days period using a rolling window approach.

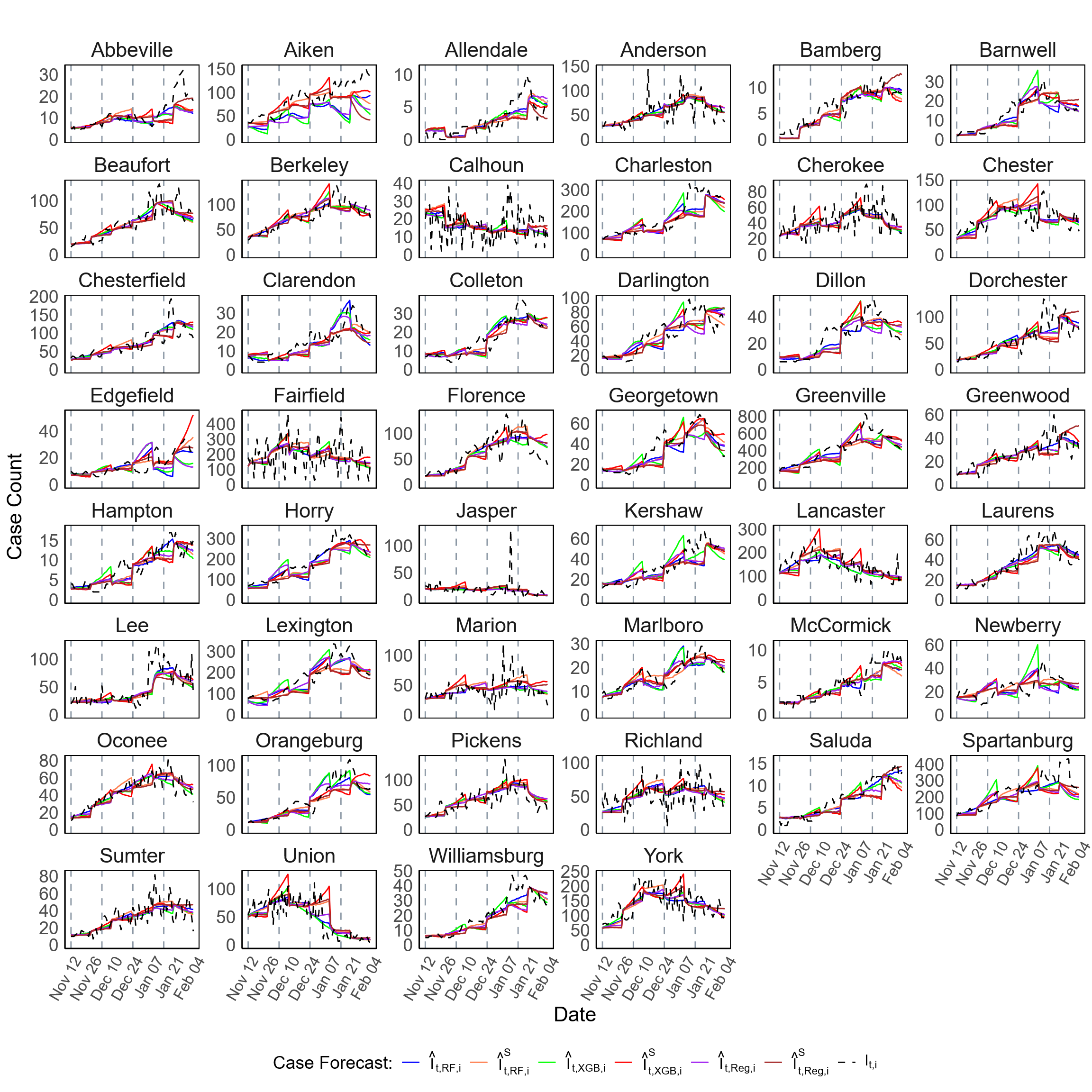

**Figure S22.** Forecast of COVID-19 case counts at the county level in SC during Scenario-1 (November 11, 2020 – February 02, 2021). This figure presents the forecast of COVID-19 case counts for all 46 counties in SC. The plots compare forecasts generated using multiple individual models against the observed daily case counts (black dashed lines, $I_{t,i}$), where $i$ represents the county and $t$ denotes the time point (day). The forecasts were generated for 14-day horizon predictions over 84 days period using a rolling window approach.

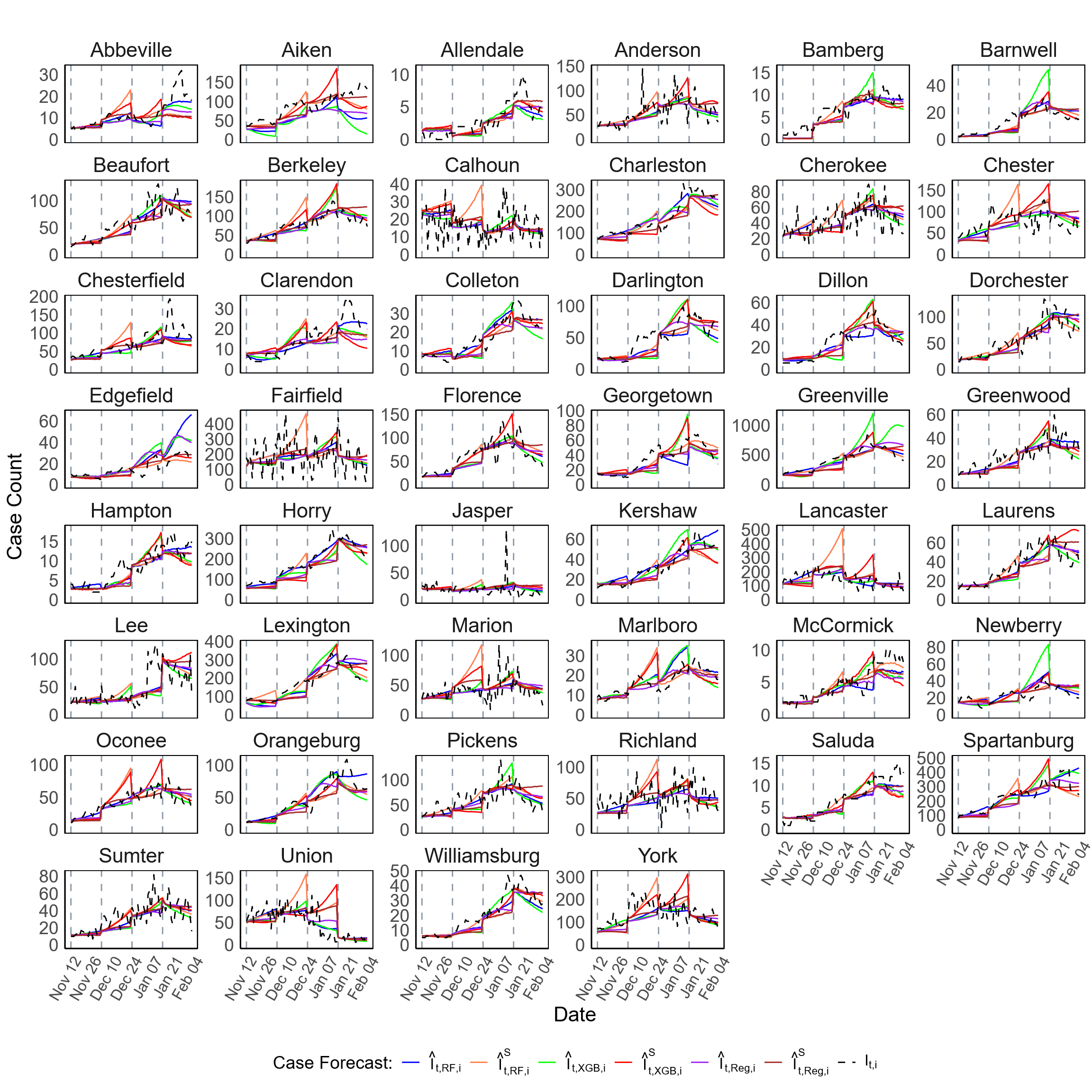

**Figure S23.** Forecast of COVID-19 case counts at the county level in SC during Scenario-1 (November 11, 2020 – February 02, 2021). This figure presents the forecast of COVID-19 case counts for all 46 counties in SC. The plots compare forecasts generated using multiple individual models against the observed daily case counts (black dashed lines, $I_{t,i}$), where $i$ represents the county and $t$ denotes the time point (day). The forecasts were generated for 21-day horizon predictions over 84 days period using a rolling window approach.

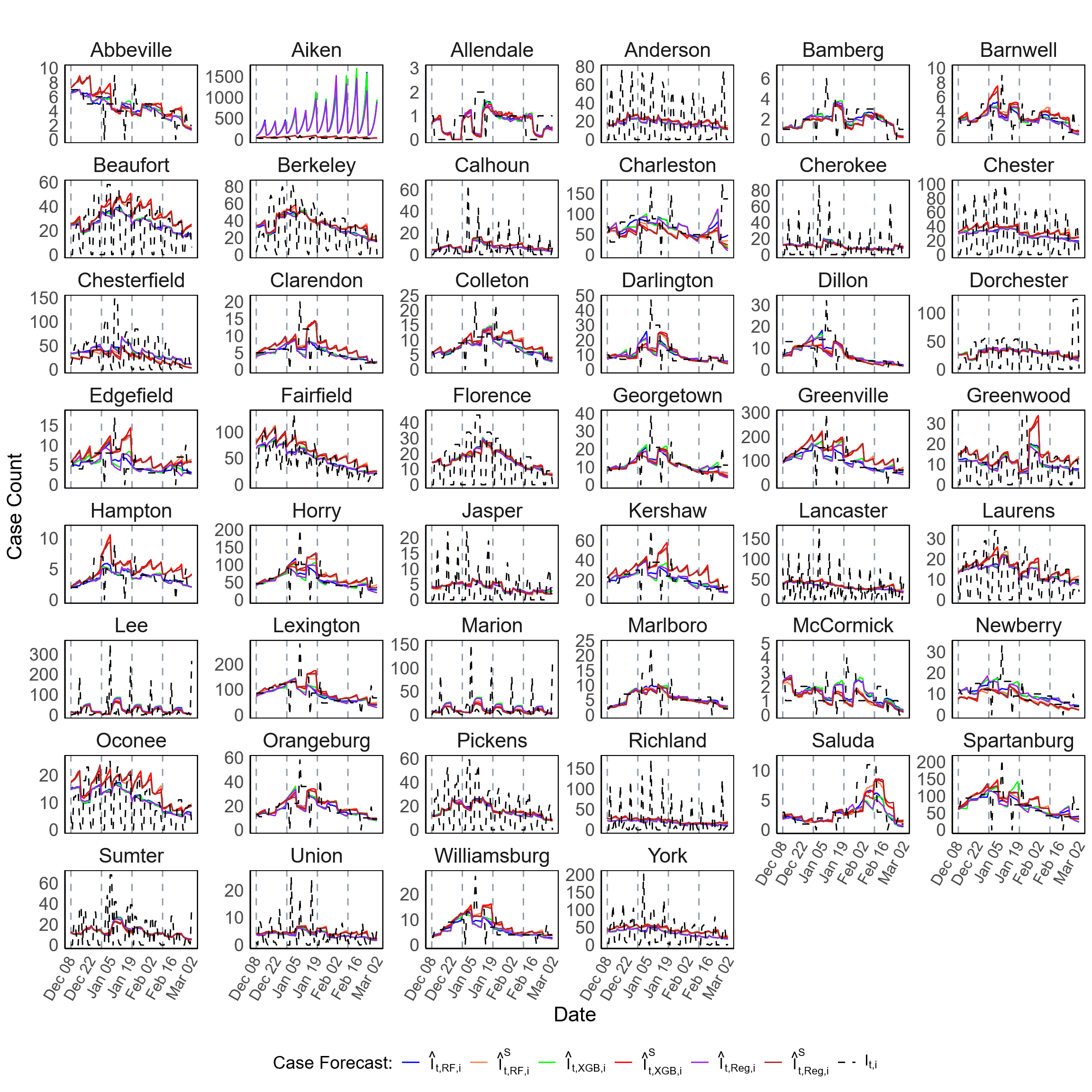

**Figure S24.** Forecast of COVID-19 case counts at the county level in SC during Scenario-2 (December 11, 2022 – March 04, 2023). This figure presents the forecast of COVID-19 case counts for all 46 counties in SC. The plots compare forecasts generated using multiple individual models against the observed daily case counts (black dashed lines, $I_{t,i}$), where $i$ represents the county and $t$ denotes the time point (day). The forecasts were generated for 7-day horizon predictions over 84 days period using a rolling window approach.

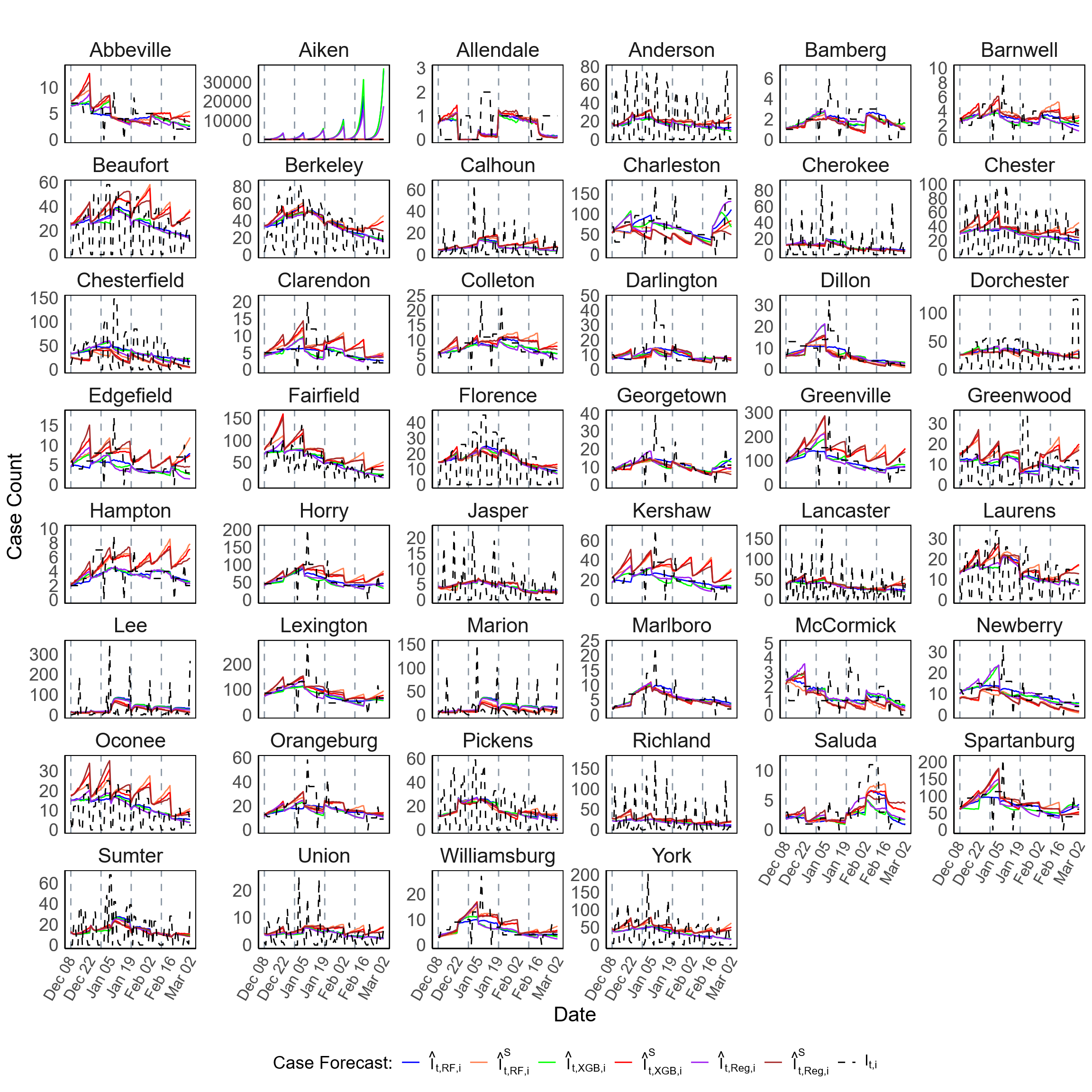

**Figure S25.** Forecast of COVID-19 case counts at the county level in SC during Scenario-2 (December 11, 2022 – March 04, 2023). This figure presents the forecast of COVID-19 case counts for all 46 counties in SC. The plots compare forecasts generated using multiple individual models against the observed daily case counts (black dashed lines, $I_{t,i}$), where $i$ represents the county and $t$ denotes the time point (day). The forecasts were generated for 14-day horizon predictions over 84 days period using a rolling window approach.

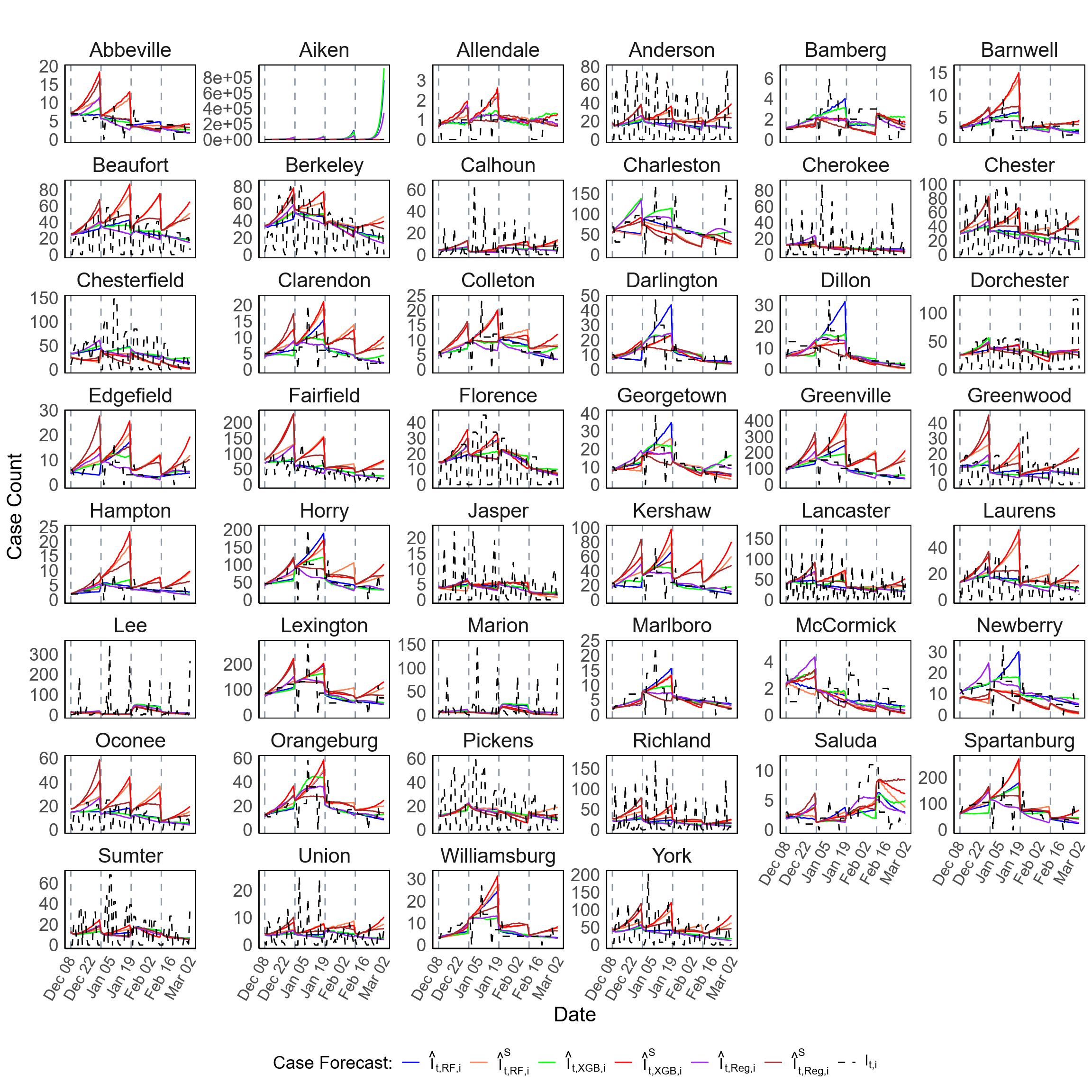

**Figure S26.** Forecast of COVID-19 case counts at the county level in SC during Scenario-2 (December 11, 2022 – March 04, 2023). This figure presents the forecast of COVID-19 case counts for all 46 counties in SC. The plots compare forecasts generated using multiple individual models against the observed daily case counts (black dashed lines, $I_{t,i}$), where $i$ represents the county and $t$ denotes the time point (day). The forecasts were generated for 21-day horizon predictions over 84 days period using a rolling window approach.
